# Cross-Cohort Generalizability of Plasma Biomarker Machine Learning Models Reveals Calibration-Driven Degradation in Clinical Utility

**DOI:** 10.64898/2026.04.09.26350514

**Authors:** Apurva Korni, Ebrahim Zandi

## Abstract

**Background:** Plasma biomarkers demonstrate strong within-cohort performance for identifying cerebral amyloid pathology, but their real-world clinical utility depends on generalization across populations and assay platforms. The impact of cross-cohort deployment on clinically actionable metrics such as negative predictive value (NPV) remains poorly characterized.

**Objective:** To evaluate the performance and portability of plasma biomarker–based machine learning models for amyloid PET prediction across independent cohorts, with emphasis on calibration and clinically relevant predictive values.

**Methods:** Data from ADNI (n=885) and A4 (n=822) were analyzed. Machine learning models were trained within each cohort to predict amyloid PET status and continuous amyloid burden (centiloids). Performance was assessed using ROC AUC, accuracy, R², and RMSE. Cross-cohort generalizability was evaluated using bidirectional transfer without retraining. Calibration, predictive values, and decision curve analysis were used to assess clinical utility.

**Results:** Within-cohort discrimination was high (AUC up to 0.913 in ADNI and 0.870 in A4), with moderate performance for centiloid prediction (R² up to 0.628 and 0.535, respectively). Cross-cohort deployment resulted in modest attenuation of AUC (∼4-7%) but substantially greater degradation in clinically actionable performance. NPV declined from 0.831 to 0.644 under ADNI→A4 transfer (∼19 percentage points) despite preserved discrimination. Calibration analyses demonstrated systematic probability misestimation, and decision curve analysis showed reduced net clinical benefit. Biomarker distribution differences across cohorts were consistent with dataset shift.

**Conclusion:** Plasma biomarker models retain discrimination across cohorts but exhibit clinically meaningful degradation in predictive value under deployment. Calibration instability and prevalence differences critically affect NPV, highlighting the need for cross-cohort validation, calibration assessment, and assay harmonization before clinical implementation.

## Introduction

Alzheimer’s disease (AD) is a major and growing public health concern. It is now well established that pathological changes in the brain begin 10-20 years before memory loss leading to clinical symptoms and diagnosis^1–3^. Despite tremendous advances in therapeutic development, the treatments available today have limited effectiveness in individuals with established symptomatic disease and substantial amyloid deposits and tau pathology^4,5–8^.

Some anti-amyloid therapeutics have demonstrated the ability to reduce amyloid plaque burden and slow clinical decline in early symptomatic AD^9, 10^. In parallel, ongoing prevention trials such as AHEAD 3-45 (NCT04468659)^11^ are testing whether similar approaches can slow biomarker progression in asymptomatic or preclinical individuals. These developments highlight the importance of identifying individuals at the earliest stages of disease, prior to the onset of cognitive symptoms, when therapeutic interventions may be most effective.

In addition to pharmacological approaches, a growing body of evidence indicates that modifiable lifestyle and vascular risk factors contribute significantly to dementia risk. Multidomain interventions targeting diet, physical activity, cognitive engagement, and cardiovascular health have demonstrated benefits on cognitive outcomes in at-risk populations, and large-scale analyses suggest that a significant proportion of dementia cases may be attributable to modifiable factors^6, 12, 13^. These findings support the concept that early identification of underlying AD pathology may provide a critical window for intervention, during which both pharmacologic and nonpharmacologic strategies can be implemented to delay or mitigate disease progression.

Accordingly, early detection of amyloid and tau pathology using blood-based biomarkers in asymptomatic individuals represents a key step toward effective prevention and disease modification. Amyloid positron emission tomography (PET) and cerebrospinal fluid (CSF) biomarkers as reference markers for identification of cerebral amyloid deposition are still accepted and widely used for reference^14–17^. However, their high cost, invasive nature, and limited accessibility restrict their broader application in screening, recruitment for clinical trials, and long-term surveillance at scale^15, 17, 18^. Due to these constraints, there has been a growing interest in the role of plasma-based biomarkers (PBBMs) as non-invasive and scalable tools for diagnosing AD pathology^16, 17, 19.^

Over the past decade, PBBMs such as Aβ42/40 ratio, phosphorylated tau species (mainly p-tau217), glial fibrillary acidic protein (GFAP), and neurofilament light chain (NfL) have demonstrated robust associations with amyloid PET positivity and neurodegeneration^19–23^. Among these, Aβ42/40 and p-tau217 have achieved the best discriminative performance for the prediction of amyloid PET. Plasma Aβ42/40 assays provided values of area under receiver operating characteristic curve (AUC) of about 0.88 for amyloid PET status and 0.94 when demographic and genetic characteristics such as age and APOE ε4 status were included^17, 24, 25^. Most recently, p-tau217 was reported to be the best PBBM that can identify asymptomatic individuals and diagnose those that have pathological underlying lesions of amyloid and tau^26, 27^. Longitudinal studies have shown that plasma p-tau217 trajectories encode temporal data regarding disease course. A recently established plasma p-tau217 “clock” model derived from repetitive biomarker measurements provides a clinically meaningful estimate of the timing of symptomatic AD emergence in harmonized cohorts^28^. These results emphasize the biological potency of plasma biomarkers, notably p-tau217, as diagnostic and diagnostic staging instruments.

However, there is an important gap in translation. Most biomarker studies, and the predictive models resulting from them, are developed and tested in a single cohort under internally reproducible assay conditions^17, 29–31^. Instead, real-world deployment depends on models being implemented across populations that vary in sample composition, disease prevalence, and importantly biomarker measurement platforms. A systematic shift in the distributions of biomarkers in cohorts can arise from differences in assay calibration, analytical methods, and handling prior to analysis^32–35^. Such changes may alter the relationship between biomarkers and disease outcomes if the basic biology remains invariable. This leads to a basic question**: do predictive models trained on plasma biomarker, genomic, and demographic data in one cohort extrapolate to another cohort in which the assay platforms and population characteristics are different?** Although machine learning (ML) methods have been adopted to combine multimodal data and attain good within-cohort performance^36–39^ the cross-cohort generalization of ML is less clear. Specifically, it remains not clear how valid and robust an individual cohort’s models (e.g., ADNI^40^) may be with respect to an independent cohort (e.g., A4^41^) which has different biomarker assays and subgroups.

This problem is particularly problematic for screening purposes in clinical practice. For preclinical populations with low disease prevalence and decisions being based on rule out, the negative predictive value (NPV), for instance, is an important performance factor^16, 17, 42^. Unlike discrimination markers such as AUC, NPV is strongly sensitive to disease prevalence and probability calibration. Even slight changes in calibration or biomarker distribution can generate significant variations in clinically actionable performance^42–45^.

In the current study, we examine performance and portability of machine learning models developed based on PBBMs under defined cross-cohort deployment contexts. We utilized PBBMs, PET, and demographics data from two complementary cohorts, ADNI^40^, which covers the full spectrum of AD clinical presentation, and A4^41^, which represents a relatively well asymptomatic population . We first establish within-cohort performance, and subsequently cross-cohort generalizability through bidirectional transfer (ADNI→A4 and A4→ADNI). We focus on discrimination, calibration, predictive values, and clinical utility. The findings indicate how variations in the PBBMs assays in different cohorts with various prevalences and clinical stages of disease affect predictive performance and highlights major hurdles of translating our PBBMs into clinical practice.

## Materials & Methods

### Study Cohorts

This study was designed as a retrospective cross-cohort validation study evaluating model portability under dataset shift conditions. We analyzed data from two independent, well-characterized cohorts: the Alzheimer’s Disease Neuroimaging Initiative (ADNI^40^) and the Anti-Amyloid Treatment in Asymptomatic Alzheimer’s Disease (A4^41^) study. ADNI is a longitudinal observational study designed to characterize the full clinical spectrum of Alzheimer’s disease, including cognitively normal individuals, participants with mild cognitive impairment (MCI), and patients with Alzheimer’s dementia^40^. The A4 study is a large secondary prevention trial focused on clinically normal, amyloid-screened older adults, representing a preclinical population with low disease prevalence^41^. Amyloid positivity prevalence was 46.5% in ADNI and 61.8% in A4.

All analyses were restricted to participants with complete data on plasma biomarkers, relevant covariates, and amyloid PET outcomes. This yielded 885 ADNI participants and 822 A4 participants with complete amyloid PET centiloid values and binary amyloid PET status. An additional 864 ADNI participants had complete clinical diagnosis data (cognitively normal, MCI, or dementia), which were not available in A4. Only baseline measurements were used for the present analyses to reflect realistic screening deployment conditions.

### Outcomes

The primary outcome was amyloid PET status, defined as amyloid positive versus negative based on established centiloid thresholds within each cohort^14^. Secondary outcomes included continuous amyloid PET burden expressed in centiloids and, in ADNI only, multiclass clinical diagnosis. These outcomes were selected to evaluate both clinically actionable classification tasks and continuous disease burden prediction.

### Plasma Biomarkers and Covariates

Plasma biomarkers and covariates datasets were acquired from ADNI and A4. The plasma biomarker assay platforms are described in ADNI^40^ and A4^41^. Biomarkers included phosphorylated tau (p-tau217), amyloid-β 40 (Aβ40), amyloid-β 42 (Aβ42), the Aβ42/40 ratio, GFAP, and NfL. In ADNI, two p-tau217 assays were available and analyzed separately to isolate assay-specific effects. All biomarker values were log-transformed where appropriate and standardized (z-scored) within cohort prior to modeling, consistent with prior plasma biomarker ML studies^38^. For transparency, cohort means and variances for primary biomarkers are provided in Supplementary Table S1 and distributional comparisons are shown in Figure 8 and Supplementary Figures S2–S4. When reporting within-cohort standardization we mean centering and scaling by each cohort’s empirical mean and standard deviation (i.e., z = (x − μ_cohort)/σ_cohort). We preserved raw-scale summary statistics (means and SDs) so that the effect of standardization versus raw-scale differences can be explicitly examined.

### Machine Learning Models

We evaluated a range of supervised machine learning models commonly used in biomarker-based prediction^38, 39^ (Table 1). Model inputs included plasma biomarkers, demographic variables, and APOEe4 status. For classification tasks (amyloid PET status and ADNI clinical diagnosis), models included logistic regression, random forest, gradient boosting (XGBoost and LightGBM), and support vector machines with radial basis function kernels. For regression tasks predicting continuous centiloid values, models included ridge regression, random forest regression, gradient boosting models (XGBoost and LightGBM), and support vector regression.

**Table 1.**
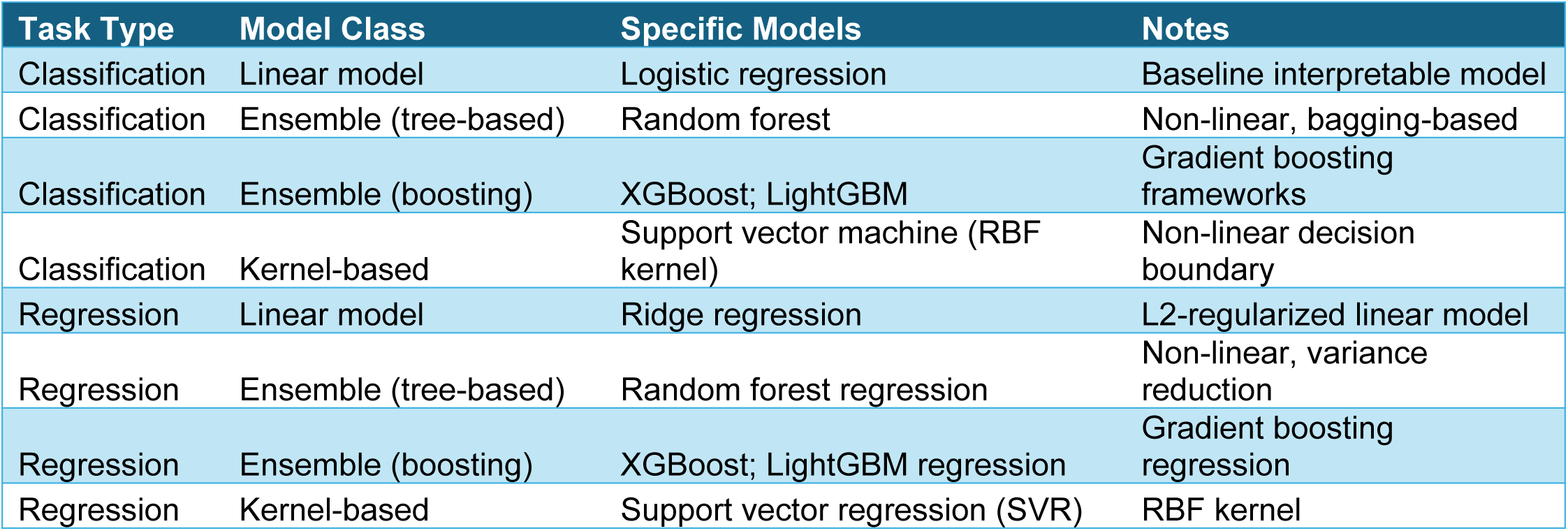
Machine learning models evaluated for classification and regression tasks.

| Task Type | Model Class | Specific Models | Notes |
| --- | --- | --- | --- |
| Classification | Linear model | Logistic regression | Baseline interpretable model |
| Classification | Ensemble (tree-based) | Random forest | Non-linear, bagging-based |
| Classification | Ensemble (boosting) | XGBoost; LightGBM | Gradient boosting frameworks |
| Classification | Kernel-based | Support vector machine (RBF kernel) | Non-linear decision boundary |
| Regression | Linear model | Ridge regression | L2-regularized linear model |
| Regression | Ensemble (tree-based) | Random forest regression | Non-linear, variance reduction |
| Regression | Ensemble (boosting) | XGBoost; LightGBM regression | Gradient boosting regression |
| Regression | Kernel-based | Support vector regression (SVR) | RBF kernel |

Linear and kernel-based models were implemented using pipelines with feature standardization, while tree-based and boosting models were trained on standardized input features without additional scaling. All models were implemented in Python using established machine learning libraries (e.g., scikit-learn, XGBoost, and LightGBM), with fixed random seeds to ensure reproducibility. Unless otherwise specified, default hyperparameters were used, consistent with prior comparative ML evaluations in neuroimaging and biomarker research^39^. Model processing, feature scaling and evaluation procedures were constant across cohorts to ensure compatibility.

### Within-Cohort Model Development and Selection

Within each cohort, models were trained and evaluated using complete-case datasets. For binary classification, discrimination was assessed using ROC AUC as the primary metric with accuracy reported as secondary metric^46^. For centiloid regression, R² was used as the primary metric with RMSE as secondary. The best-performing model for each cohort and outcome was selected based on the primary metric and carried forward as the reference model for cross-cohort validation.

### Cross-Cohort Validation Strategy

To assess model generalizability, we performed bidirectional cross-cohort validation (A4→ADNI and ADNI→A4) consistent with recent work on dataset shift and model robustness^47^. Models were trained on all eligible participants in the source cohort and applied without retraining to the target cohort. Separate cross-cohort models were trained for each p-tau217 assay available in ADNI to isolate assay-specific transfer effects. No hyperparameter tuning or recalibration was performed during initial cross-cohort testing unless explicitly noted.

### Calibration, Predictive Values, and Sensitivity Analyses

Given the clinical importance of rule-out performance in preclinical populations, NPV was evaluated as a key secondary metric^17, 48^. PPV and NPV were estimated at fixed sensitivity thresholds defined in the training cohort using Youden’s index and applied unchanged to the test cohort to simulate deployment; confidence intervals were obtained by bootstrap resampling (1,000 iterations)^49^. Calibration was assessed using Brier scores, calibration slope, and intercept statistics^43, 45, 47^, and we considered modern ML calibration behavior and practical recalibration approaches in our supplementary analyses. Sensitivity analyses included reciprocal training–testing directions, algorithm comparisons, p-tau217 assay variants, and subgroup analyses by APOE ε4 carrier status and sex.

### Statistical Analysis and Reproducibility

All analyses were conducted using version-controlled Python and R scripts with pre-specified analysis plans to minimize analytic bias and data leakage. The study adhered to STROBE and TRIPOD reporting recommendations where applicable^50, 51^. Summary statistics are reported with appropriate confidence intervals.

### Ethics Statement

All data were obtained from ADNI and A4 under approved protocols with informed consent from all participants and institutional review board approval at participating sites.

## RESULTS

### Study Design and Population

The overall study design and analytic framework are shown in Figure 1. The analysis included two complementary cohorts, ADNI and A4, with within-cohort model development and internal validation followed by bidirectional cross-cohort deployment (ADNI→A4 and A4→ADNI). This design enabled evaluation of both internal performance and portability across distinct clinical and prevalence contexts. The analytic strategy, including cohort composition, outcome definitions, and performance metrics, is summarized in Figure 1. Consistent with prior work highlighting the importance of dataset shift in clinical machine learning, the cross-cohort design was intended to stress-test portability under realistic deployment conditions^47^.

**Figure 1.**
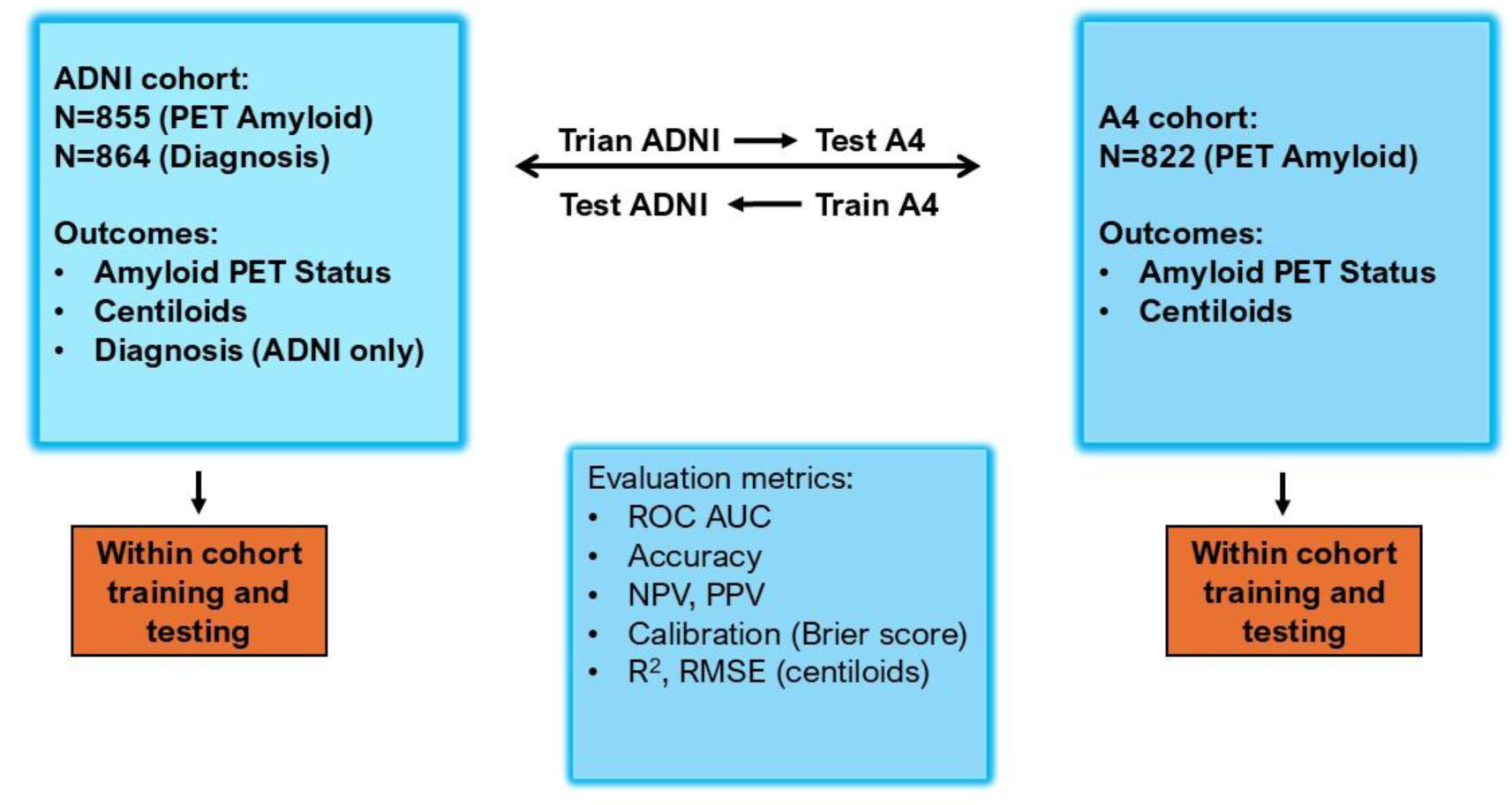
Study design and bidirectional cross-cohort validation framework. Schematic overview of cohort composition, modeling strategy, and evaluation metrics. The ADNI cohort included N=855 participants with amyloid PET data and N=864 with clinical diagnosis; outcomes included amyloid PET status, continuous centiloid values, and diagnosis (ADNI only). The A4 cohort included N=822 participants with amyloid PET data; outcomes included amyloid PET status and centiloids. Within-cohort training and testing were performed separately in ADNI and A4. Cross-cohort generalizability was evaluated using bidirectional transfer (ADNI→A4 and A4→ADNI) without retraining. Model performance was assessed using discrimination metrics (ROC AUC, accuracy), predictive values (NPV, PPV), calibration (Brier score), and regression performance metrics (R2 and RMSE for centiloids).

A total of 1,707 participants were included after restricting analyses to individuals with complete plasma biomarker, covariate, and amyloid PET data (ADNI, n = 885; A4, n = 822). ADNI spanned the full clinical spectrum, including cognitively normal individuals, participants with mild cognitive impairment, and patients with Alzheimer dementia^40^. In contrast, A4 comprised cognitively normal individuals enriched for amyloid negativity^41^. Amyloid positivity prevalence was 46.5% in ADNI and 61.8% in A4, establishing distinct clinical and prevalence settings for evaluating model portability^26^.

### Within-Cohort Prediction of Amyloid PET Status and Clinical Outcomes

Within-cohort supervised machine learning models demonstrated strong performance for amyloid PET classification in both cohorts (Table 2; Figure 2). In ADNI, random forest achieved the highest discrimination for amyloid status (ROC AUC = 0.913; accuracy = 0.845), with comparable performance observed across gradient boosting models and support vector machines (AUC range: 0.905–0.913). Logistic regression also performed well (AUC = 0.900), indicating that linear models captured a substantial portion of the underlying biological signal. These findings are consistent with prior reports showing strong plasma p-tau217 and plasma amyloid biomarker discrimination for amyloid PET status^21, 26, 52, 53^.

**Figure 2.**
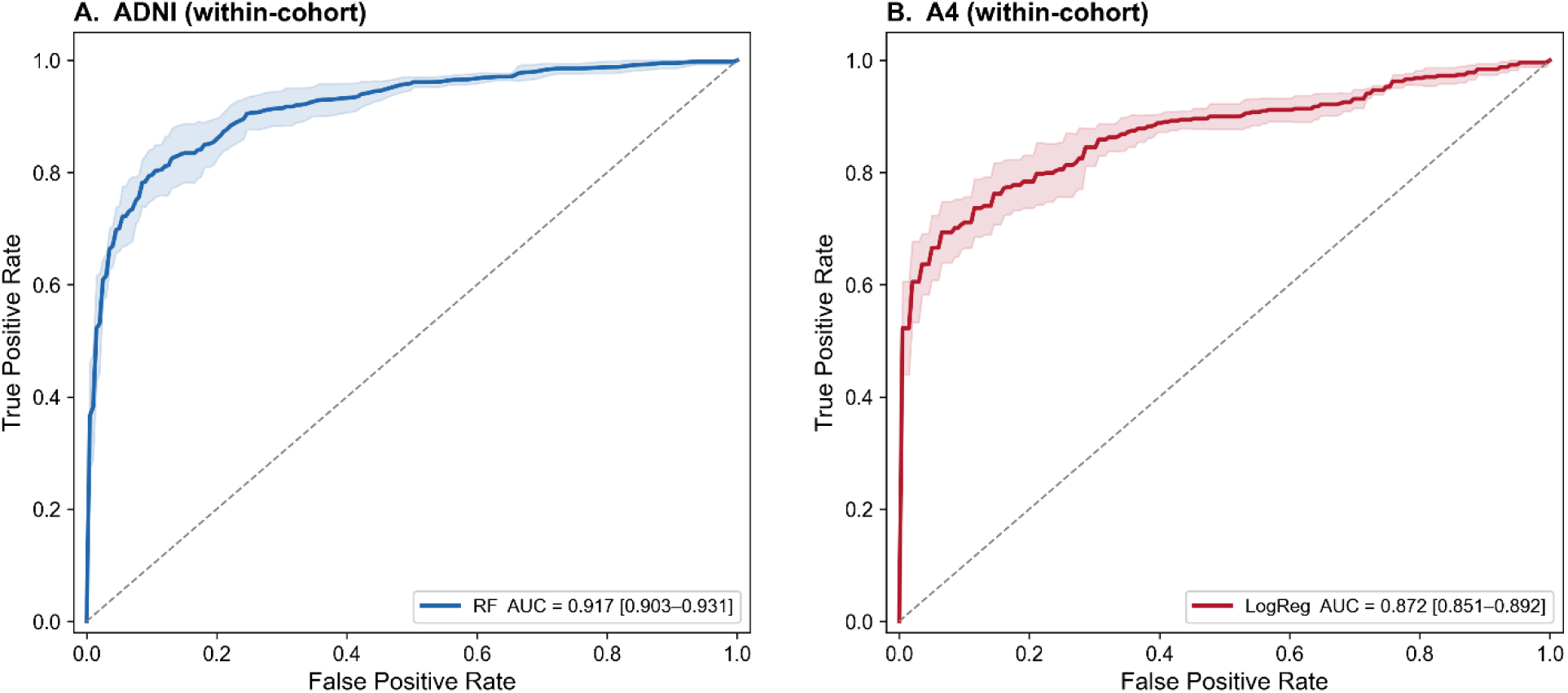
Receiver operating characteristic (ROC) curves for within-cohort prediction models. (A) ROC curve for the random forest (RF) model evaluated within the ADNI cohort. (B) ROC curve for the logistic regression (LogReg) model evaluated within the A4 cohort. In both panels, the solid line represents the mean model performance and the shaded region indicates the 95% confidence interval. The diagonal dashed line denotes chance-level performance. Model discrimination was higher for the RF model in ADNI (AUC = 0.917, 95% CI: 0.903–0.931) compared to the logistic regression model in A4 (AUC = 0.872, 95% CI: 0.851–0.892).

**Table 2.**
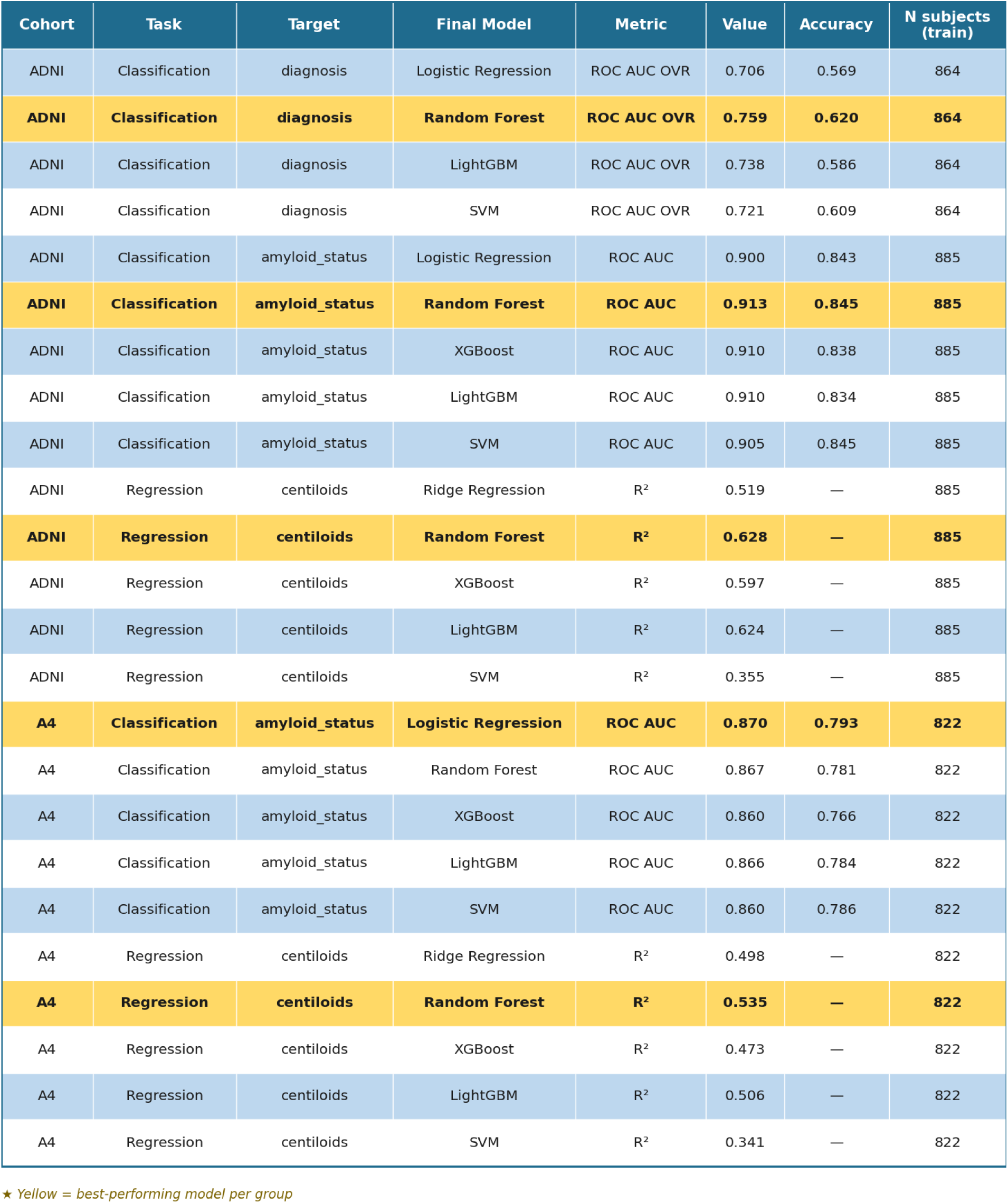
Within cohort ML models.

In A4, logistic regression achieved the highest performance for amyloid classification (ROC AUC = 0.870; accuracy = 0.793), with tree-based and kernel-based models showing similar performance (AUC range: 0.860–0.867). Discrimination was modestly lower in A4 than in ADNI, consistent with the narrower biologic dynamic range and lower clinical heterogeneity of the preclinical cohort^21, 54–58^.

Within ADNI, multiclass classification of clinical diagnosis yielded lower discrimination than amyloid classification, with the best performance observed for random forest (ROC AUC = 0.759). This likely reflects the more complex relationship between plasma biomarkers and clinically defined stages of cognitive impairment.

### Within-Cohort Prediction of Continuous Amyloid Burden

Models predicting continuous amyloid PET burden, expressed as centiloids, also performed well within cohort (Table 2). In ADNI, random forest provided the best performance (R^2^ = 0.628), followed closely by LightGBM (R^2^ = 0.624) and XGBoost (R^2^ = 0.597). In A4, performance was more modest but still meaningful, with the best result achieved by random forest (R^2^ = 0.535). These findings suggest that plasma biomarkers capture not only binary amyloid status but also a substantial portion of quantitative amyloid burden^19, 31, 58^.

Across both cohorts, non-linear ensemble methods consistently outperformed linear regression models, indicating that the relationship between plasma biomarkers and amyloid burden is at least partly non-linear.

### Model Structure and Feature Contributions

To further characterize model structure and biological coherence, we examined standardized logistic regression coefficients (Supplementary Figure S1), feature importance rankings (Supplementary Figure S2), and SHAP-based interpretability analyses (Supplementary Figure S3). Standardized logistic regression coefficients showed that p-tau217 made the largest contribution to amyloid classification in both cohorts, with APOE ε4 allele count and the Aβ42/40 ratio also demonstrating significant effects. Feature importance rankings across random forest, XGBoost, and LightGBM models consistently identified p-tau217 as the most influential predictor, followed by APOE ε4 and Aβ42/40.

SHAP analyses further showed that higher p-tau217 values increased the predicted probability of amyloid positivity, whereas higher Aβ42/40 values decreased risk. These effects were monotonic and directionally consistent across cohorts. Together, these analyses indicate that biological signal structure was stable across modeling approaches within each cohort and that predictive performance was driven primarily by the biomarker signal rather than model architecture.

### Cross-Cohort Generalizability of Amyloid PET Classification

Building on the strong within-cohort performance observed in Table 2 and Figure 3, we next evaluated model generalizability under cross-cohort deployment (Table 3; Figure 3). Table 3 highlights a dissociation between preserved discrimination and degradation of clinically actionable metrics.

**Figure 3.**
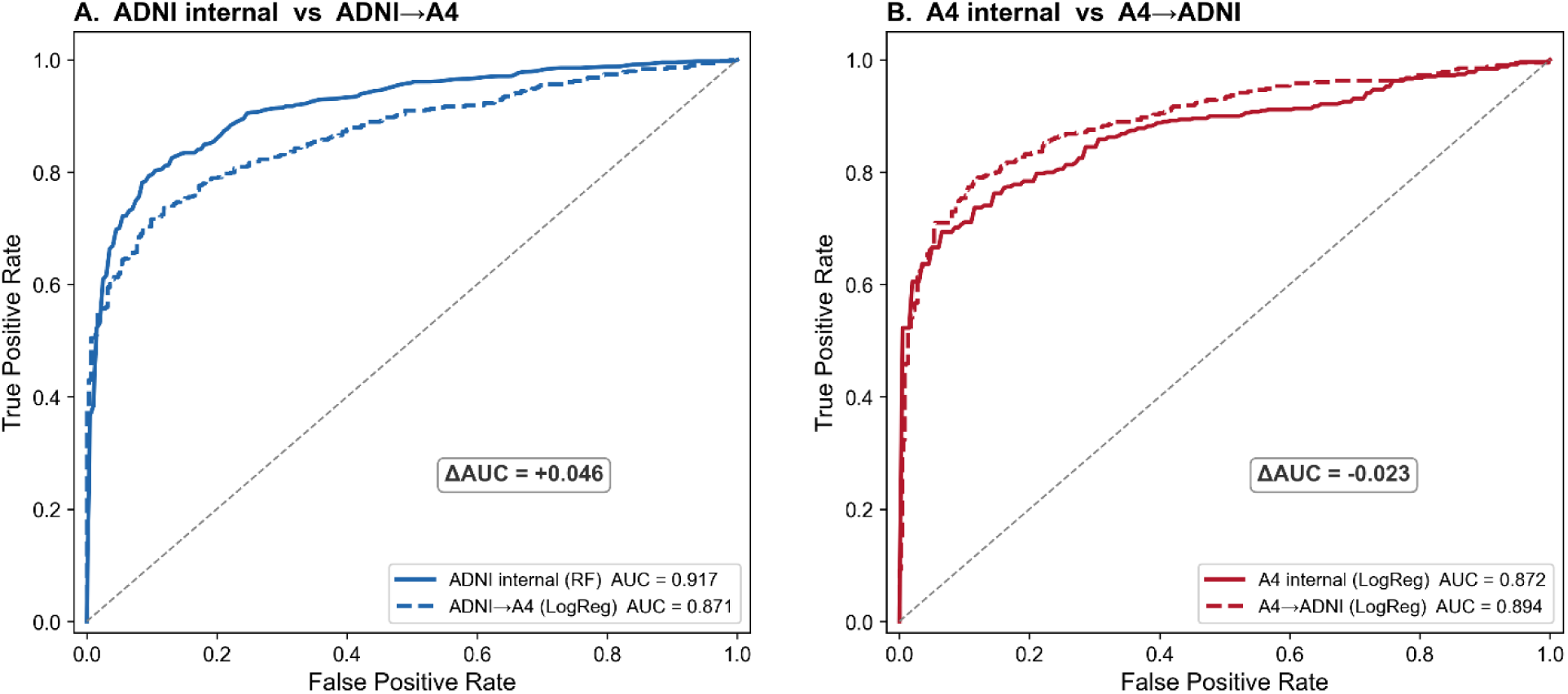
Comparison of within-cohort and cross-cohort model performance. (A) ROC curves showing performance of the model trained and evaluated within the ADNI cohort (solid line) alongside its performance when applied to the A4 cohort (dashed line). (B) ROC curves showing performance of the model trained and evaluated within the A4 cohort (solid line) and its application to the ADNI cohort (dashed line). The diagonal dashed line represents chance-level classification. In panel A, performance declined when the model was transferred from ADNI to A4 (AUC = 0.917 vs. 0.871; ΔAUC = 0.046). In panel B, the model trained in A4 showed slightly improved performance when applied to ADNI (AUC = 0.872 vs. 0.894; ΔAUC = −0.023), suggesting differences in generalizability between cohorts.

**Table 3.** Cross-Cohort Generalizability of Amyloid PET Classification.

| direction | model | n_test | ppv | npv | accuracy | AUC | f1 | is_best |
| --- | --- | --- | --- | --- | --- | --- | --- | --- |
| ADNI->A4 | LogReg | 822 | 0.930851 | 0.643498 | 0.774939173 | 0.870824 | 0.79096 | TRUE |
| ADNI->A4 | RF | 822 | 0.945559 | 0.621564 | 0.759124088 | 0.868162 | 0.769231 | FALSE |
| ADNI->A4 | XGB | 822 | 0.919668 | 0.616052 | 0.749391727 | 0.842936 | 0.763218 | FALSE |
| ADNI->A4 | LGBM | 822 | 0.912568 | 0.616228 | 0.748175182 | 0.845302 | 0.763429 | FALSE |
| ADNI->A4 | SVM | 822 | 0.94152 | 0.610417 | 0.748175182 | 0.864258 | 0.756757 | FALSE |
| A4->ADNI | LogReg | 885 | 0.736082 | 0.865 | 0.794350282 | 0.894217 | 0.796875 | TRUE |
| A4->ADNI | RF | 885 | 0.736952 | 0.857143 | 0.792090395 | 0.886782 | 0.793258 | FALSE |
| A4->ADNI | XGB | 885 | 0.718941 | 0.852792 | 0.778531073 | 0.881723 | 0.782705 | FALSE |
| A4->ADNI | LGBM | 885 | 0.718941 | 0.852792 | 0.778531073 | 0.884048 | 0.782705 | FALSE |
| A4->ADNI | SVM | 885 | 0.69505 | 0.842105 | 0.75819209 | 0.873356 | 0.766376 | FALSE |
| ADNI->A4 | LogReg | 822 | 0.869347 | 0.615566 | 0.738442822 | 0.83465 | 0.762955 | FALSE |
| ADNI->A4 | RF | 822 | 0.869266 | 0.663212 | 0.772506083 | 0.854479 | 0.802116 | TRUE |
| ADNI->A4 | XGB | 822 | 0.853774 | 0.630653 | 0.745742092 | 0.823616 | 0.775991 | FALSE |
| ADNI->A4 | LGBM | 822 | 0.86014 | 0.643766 | 0.756690998 | 0.834933 | 0.78678 | FALSE |
| ADNI->A4 | SVM | 822 | 0.903315 | 0.604348 | 0.736009732 | 0.830282 | 0.750861 | FALSE |
| A4->ADNI | LogReg | 885 | 0.799523 | 0.83691 | 0.81920904 | 0.882473 | 0.807229 | TRUE |
| A4->ADNI | RF | 885 | 0.756579 | 0.846154 | 0.8 | 0.880694 | 0.795848 | FALSE |
| A4->ADNI | XGB | 885 | 0.75388 | 0.836406 | 0.794350282 | 0.878902 | 0.788863 | FALSE |
| A4->ADNI | LGBM | 885 | 0.716327 | 0.848101 | 0.775141243 | 0.881882 | 0.779134 | FALSE |
| A4->ADNI | SVM | 885 | 0.703252 | 0.834606 | 0.761581921 | 0.86658 | 0.766334 | FALSE |

When models trained in A4 were applied to ADNI, discrimination remained high, with ROC AUC values ranging from approximately 0.87 to 0.89 across models. The best performance in this direction was observed for logistic regression (AUC = 0.894; accuracy = 0.794). Corresponding NPV values remained relatively high (approximately 0.84–0.87), consistent with the clinical characteristics of ADNI.

In contrast, when models trained in ADNI were applied to A4, discrimination declined only modestly, with AUC values of approximately 0.83–0.87 across models, but clinically actionable performance deteriorated substantially. In this direction, NPV declined to approximately 0.60–0.66 across models, despite AUC remaining relatively high.

Accuracy and F1 score also decreased modestly, with accuracy ranging from approximately 0.74 to 0.77 and F1 score from approximately 0.75 to 0.79. This pattern demonstrates that models with similar discrimination can produce markedly different classification performance depending on the deployment cohort.

Relative to internal ADNI validation (AUC = 0.913), transfer to A4 produced an absolute AUC reduction of approximately 4% to 7%. Although modest in magnitude, this reduction was consistent across algorithms and biomarker configurations, indicating systematic performance attenuation under dataset shift^47^. Importantly, predictor rankings remained stable under transfer (Supplementary Figures S2 and S3), suggesting that the decline reflects distributional and calibration differences rather than loss of the underlying biological signal.

These findings highlight a gap between internal validation and real-world deployment conditions. In particular, the direction-dependent decline in NPV underscores the sensitivity of screening-relevant metrics to cohort differences and demonstrates that discrimination alone is insufficient to evaluate clinical utility^17, 48^.

### Cross-Cohort Prediction of Continuous Amyloid Burden

We next evaluated plasma biomarker–based prediction of continuous amyloid PET burden under cross-cohort deployment (Figure 4). Overall, centiloid prediction performance was substantially attenuated relative to within-cohort results. Across both transfer directions, cross-cohort models achieved R^2^ values ranging from approximately 0.35 to 0.43, with RMSE values between 31.0 and 36.6 centiloids, depending on model and feature set.

**Figure 4.**
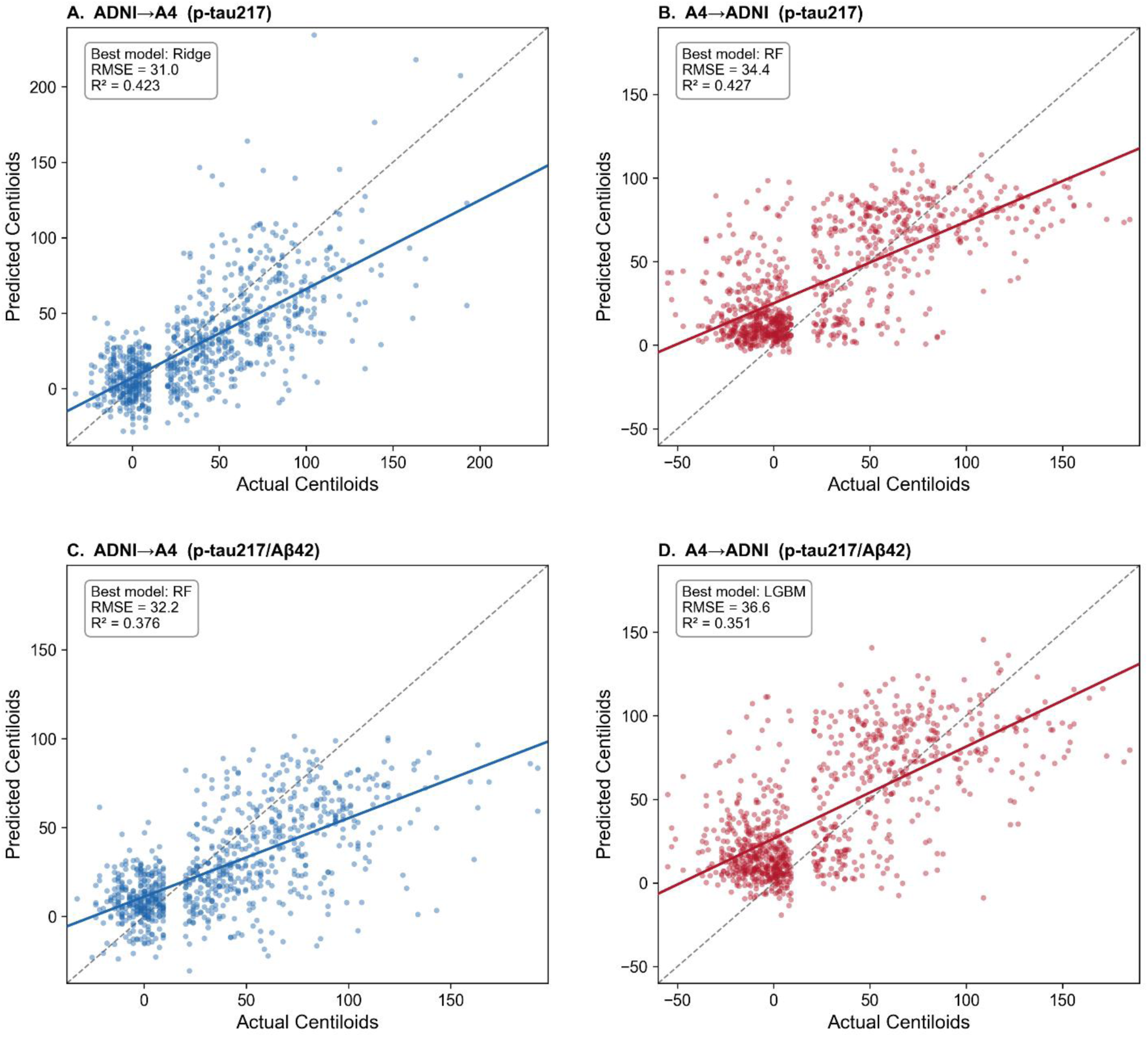
Cross-cohort prediction of plasma p-tau217 and p-tau217/Aβ42. Scatterplots show predicted versus observed Centiloid values for models trained in one cohort and evaluated in the other. (A) ADNI→A4 prediction of p-tau217 using a ridge regression model (RMSE = 31.0, R² = 0.423). (B) A4→ADNI prediction of p-tau217 using a random forest model (RMSE = 34.4, R² = 0.427). (C) ADNI→A4 prediction of p-tau217/Aβ42 using a random forest model (RMSE = 32.2, R^2^ = 0.376). (D) A4→ADNI prediction of p-tau217/Aβ42 using a light gradient boosting machine (LGBM) model (RMSE = 36.6, R^2^ = 0.351). Each point represents an individual participant. Solid lines indicate fitted regression lines, and dashed lines represent the line of identity (y = x). Across panels, models show moderate correspondence between predicted and observed Centiloid values, with some dispersion at higher values.

This represents a marked reduction from within-cohort performance (ADNI R^2^ = 0.628; A4 R^2^ = 0.535), indicating that continuous prediction is more sensitive to cross-cohort differences than binary classification^47^.

When models trained in ADNI were applied to A4 (Figure 4A and 4C), the best-performing models achieved R^2^ = 0.423 and RMSE = 31.0 for p-tau217 alone using ridge regression, and R^2^ = 0.376 with RMSE = 32.2 for p-tau217 plus Aβ42 using random forest. Scatter plots showed substantial dispersion around the identity line, particularly at higher centiloid values, and predicted values systematically underestimated higher amyloid burden.

When models trained in A4 were applied to ADNI (Figure 4B and 4D), performance was similarly attenuated. The best results in this direction were R² = 0.427 and RMSE = 34.4 for p-tau217 alone using random forest, and R² = 0.351 with RMSE = 36.6 for p-tau217 plus Aβ42 using LightGBM. Predictions showed greater dispersion across the full centiloid range, with particularly large variance at intermediate and high values. In this direction, both underestimation and overestimation were observed, indicating instability in the mapping between biomarker values and amyloid burden.

In both transfer directions, inclusion of additional biomarkers did not improve performance and in some cases yielded lower R^2^ values. This suggests that increased model complexity did not mitigate cross-cohort degradation and may have increased sensitivity to distributional differences.

Across all panels, predicted values showed greater scatter around the identity line, reduced slope relative to the ideal, and compression at higher centiloid values. These patterns indicate that models trained in one cohort did not fully capture the dynamic range of amyloid burden in the other cohort.

### Clinical Impact: Negative Predictive Value Under Cross-Cohort Deployment

To directly quantify the impact of cross-cohort deployment on clinically actionable performance, we evaluated negative predictive value across training-testing directions (Figure 5). Within-cohort models showed high NPV in both datasets. In ADNI, NPV was 0.831 (95% CI, 0.799–0.863), whereas in A4 it was 0.701 (95% CI, 0.655–0.750). The higher NPV in ADNI is consistent with its lower amyloid prevalence and broader disease spectrum, which favors rule-out performance^17, 42, 43, 59 48^.

**Figure 5.**
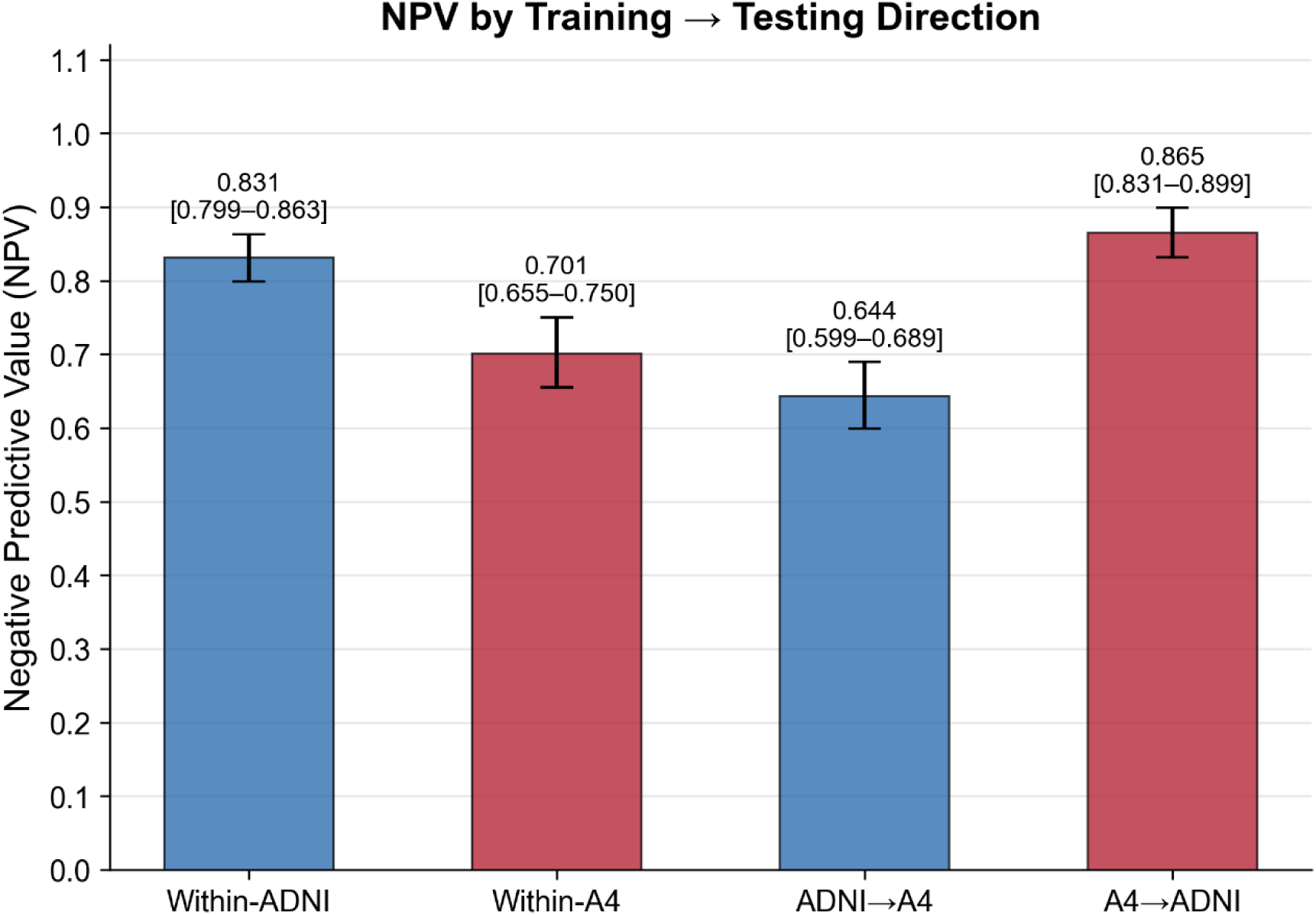
Negative predictive value (NPV) across training and testing scenarios. Bar plot showing NPV for models evaluated within-cohort (ADNI and A4) and across cohorts (ADNI→A4 and A4→ADNI). Within-cohort performance was higher in ADNI (NPV = 0.831, 95% CI: 0.799–0.863) than in A4 (NPV = 0.701, 95% CI: 0.655–0.750). When models were applied across cohorts, NPV decreased for ADNI→A4 (0.644, 95% CI: 0.599–0.689) but increased for A4→ADNI (0.865, 95% CI: 0.831–0.899). Error bars represent 95% confidence intervals.

Under cross-cohort deployment, NPV showed substantial and direction-dependent degradation. When ADNI-trained models were applied to A4, NPV declined to 0.644 (95% CI, 0.599–0.689), representing an absolute reduction of approximately 19 percentage points relative to internal ADNI performance and approximately 6 percentage points relative to internal A4 performance. In contrast, when A4-trained models were applied to ADNI, NPV increased to 0.865 (95% CI, 0.831–0.899), exceeding both within-cohort values. This asymmetry reflects the interaction between model sensitivity and cohort-specific prevalence.

These results indicate that NPV depends strongly on both the training cohort and the deployment population. The pronounced reduction in NPV under ADNI→A4 transfer suggests that models trained in clinically heterogeneous populations may not generalize well to preclinical settings, where rule-out performance is especially important.

Conversely, the increase in NPV under A4→ADNI transfer reflects the interaction of higher sensitivity with lower prevalence in the target cohort and should not be interpreted as improved calibration or discrimination.

These findings reveal a clear dissociation between discrimination and clinical utility. Although ROC AUC remained relatively stable across cohorts, NPV varied substantially under cross-cohort deployment. This indicates that screening-relevant performance cannot be inferred reliably from discrimination alone.

### Calibration Shift Under Cross-Cohort Deployment

To evaluate the reliability of predicted probabilities under cross-cohort deployment, we examined calibration curves for models trained in one cohort and applied to another, compared with within-cohort calibration (Figure 6). When models trained in ADNI were applied to A4, predicted probabilities showed systematic miscalibration relative to observed amyloid positivity rates. The calibration curve deviated from the identity line across the probability range, indicating that predicted risks were not aligned with observed frequencies.

**Figure 6.**
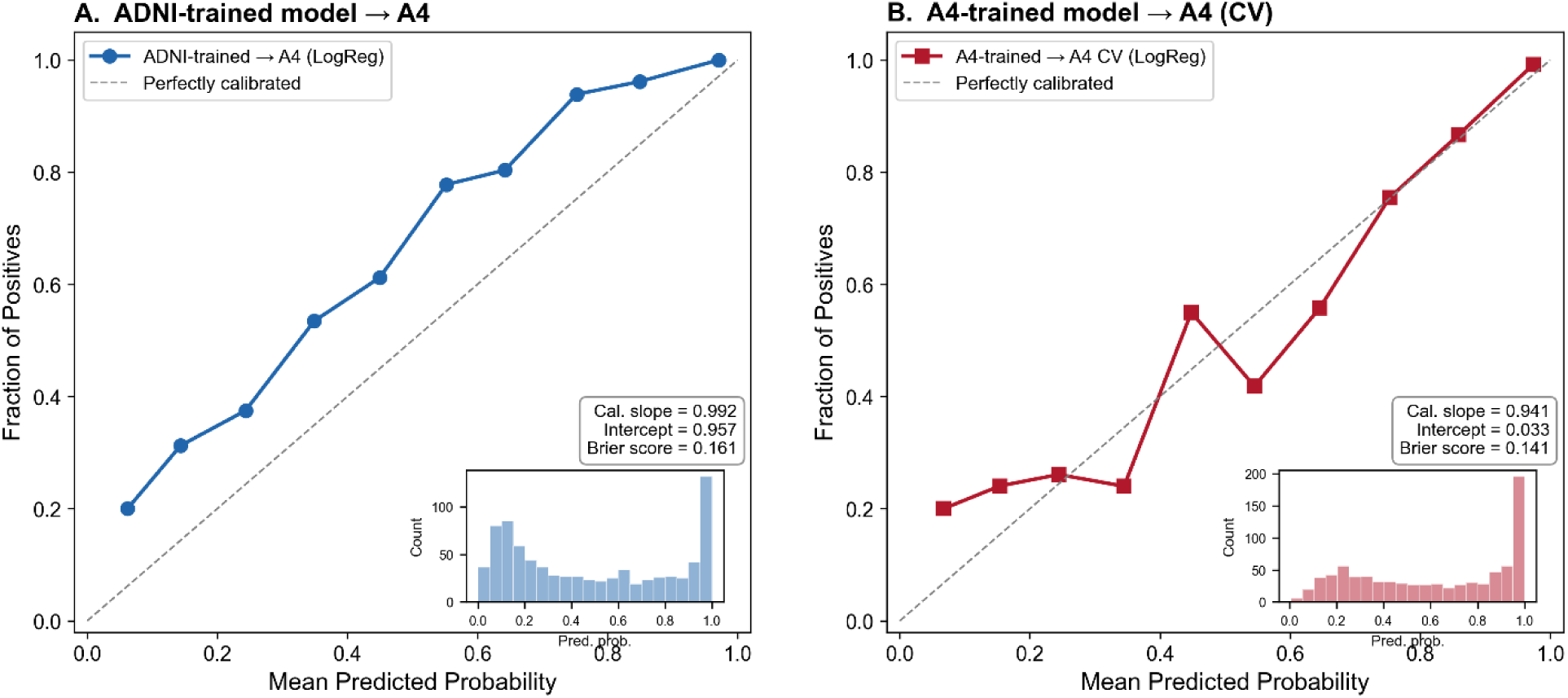
Calibration of predicted probabilities in external and internal validation. Calibration plots compare predicted probabilities with observed outcomes in the A4 cohort. (A) Performance of the model trained in ADNI and applied to A4. (B) Performance of the model trained and cross-validated within A4. The x-axis shows mean predicted probability, and the y-axis shows the observed fraction of positives within each bin. The dashed diagonal line indicates perfect calibration. In panel A, predictions from the ADNI-trained model were generally well calibrated in A4 (calibration slope = 0.992, intercept = 0.957; Brier score = 0.161). In panel B, the A4-trained model also showed good calibration under cross-validation (slope = 0.941, intercept = 0.033; Brier score = 0.141), with some deviation at intermediate probability ranges. Insets display the distribution of predicted probabilities for each model.

This miscalibration was most pronounced in the low-to-intermediate probability range, where observed event rates exceeded predicted probabilities, indicating underestimation of risk. At higher predicted probabilities, the curve approached the identity line but remained imperfect. Calibration slope and intercept values (slope = 0.992; intercept = 0.957) indicated a substantial shift in baseline probability calibration, and the Brier score was elevated (0.161), reflecting reduced probabilistic accuracy^43, 45, 47^.

In contrast, within-cohort A4 models demonstrated near-ideal calibration, with the curve closely following the identity line across the full probability range. Calibration metrics were improved relative to cross-cohort deployment (slope = 0.941; intercept = 0.033; Brier score = 0.141), supporting better probabilistic accuracy under internal validation^43, 45, 47^.

These findings show that preserved discrimination does not imply preserved calibration. Despite relatively stable AUC under transfer, predicted probabilities became systematically biased, producing misaligned risk estimates. This calibration shift provides a direct mechanistic explanation for the observed decline in NPV. Because NPV depends on accurate probability estimates and threshold selection, even modest miscalibration can meaningfully alter rule-out performance.

### Biomarker Distribution Shift as a Mechanistic Contributor

To investigate the origin of cross-cohort performance degradation, we examined raw and standardized biomarker distributions across ADNI and A4 (Figure 7). For p-tau217, ADNI showed a substantially higher mean than A4 (0.363 vs 0.173; Δ = 0.189), along with a broader right-skewed tail, indicating greater variance and dynamic range. For the Aβ42/40 ratio, the difference was smaller but still consistent (0.0576 in ADNI vs 0.0929 in A4; Δ = −0.0353), and the two cohort distributions remained clearly separated.

**Figure 7.**
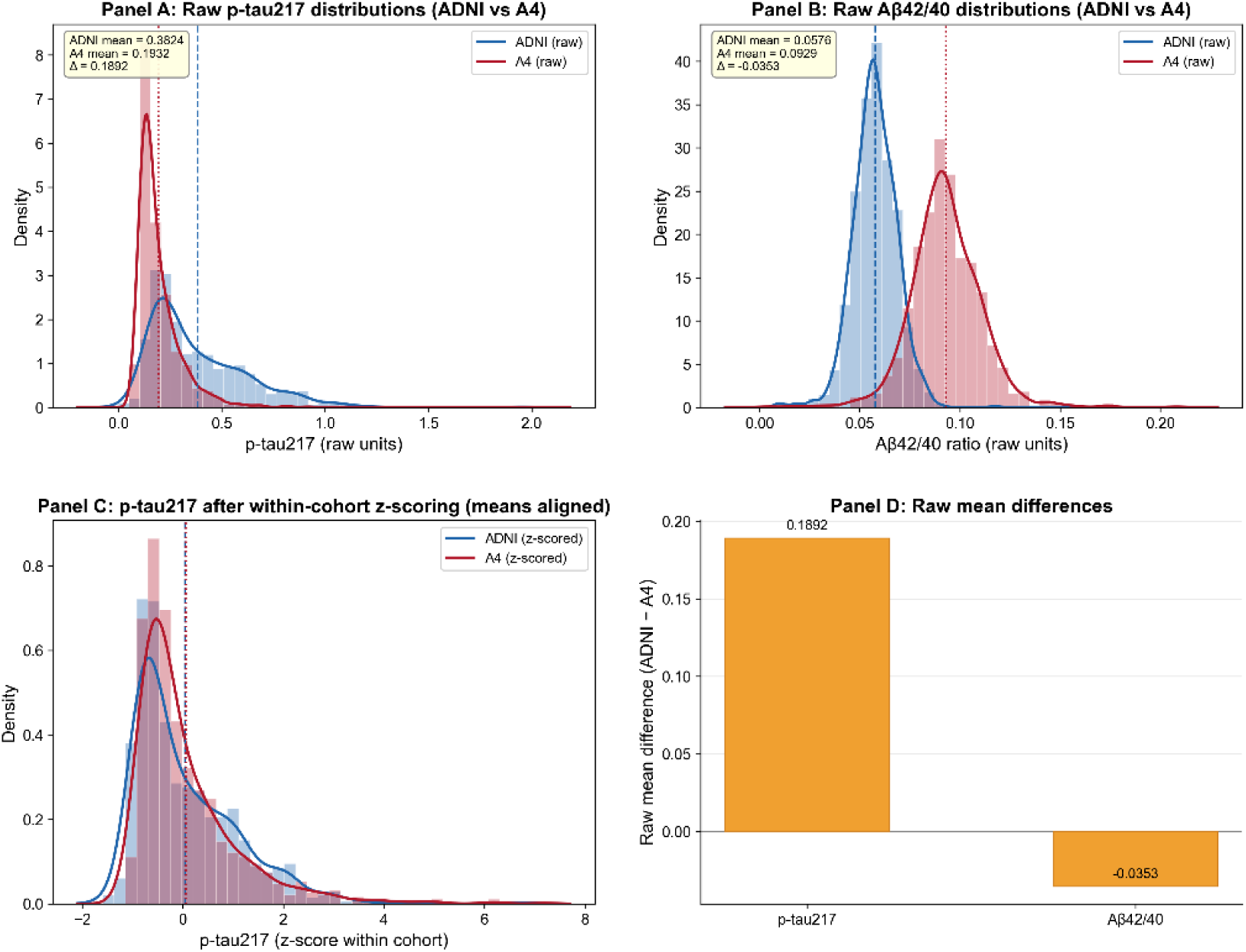
Biomarker distribution shift between ADNI and A4 cohorts. (A) Raw p-tau217 distributions (histogram + kernel density estimate) in ADNI (blue) and A4 (green). Vertical lines indicate cohort means; inset reports means and their difference (Δ). (B) Raw Aβ42/40 distributions (histogram + KDE) with cohort means and Δ annotated. (C) p-tau217 distributions after within-cohort z-scoring (means aligned to zero), illustrating that standardization masks cohort mean shifts while leaving distributional shape and variance differences intact. (D) Bar plot quantifying the raw mean differences (ADNI mean − A4 mean) for both biomarkers. These panels together illustrate how cohort-specific biomarker distributions can induce dataset shift that is not fully addressed by simple within-cohort standardization.

**Figure 8.**
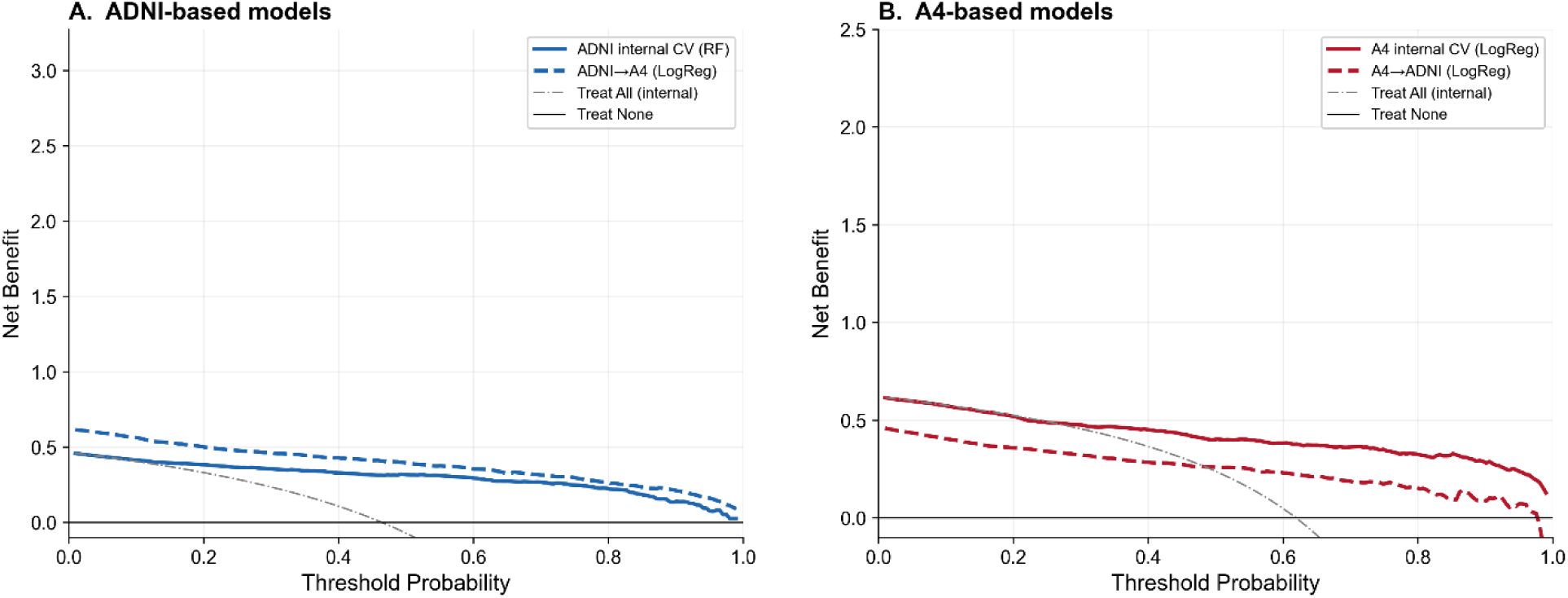
Decision curve analysis comparing within-cohort and cross-cohort model deployment. Decision curve analysis (DCA) was performed to evaluate the clinical net benefit of plasma biomarker–based machine learning models across a range of threshold probabilities. Net benefit is plotted as a function of the probability threshold used to classify amyloid PET positivity. Panel A: ADNI-based models. Solid blue line represents ADNI internal cross-validation performance (random forest). Dashed blue line represents the ADNI-trained model applied to A4 (ADNI→A4; logistic regression). The gray dash-dotted line indicates the “treat all” strategy under internal ADNI prevalence. The horizontal black line represents “treat none.” Panel B: A4-based models. Solid red line represents A4 internal cross-validation performance (logistic regression). Dashed red line represents the A4-trained model applied to ADNI (A4→ADNI). The gray dash-dotted line indicates the “treat all” strategy under internal A4 prevalence. The black horizontal line represents “treat none.” Net benefit was calculated using standard decision curve methodology, incorporating the relative weighting of false positives and false negatives determined by the chosen probability threshold. Curves above both “treat all” and “treat none” indicate clinically useful decision-making across that threshold range.

Within-cohort z-scoring aligned cohort means to zero, but differences in distribution shape, variance, and skewness persisted. In particular, the ADNI p-tau217 distribution retained a heavier right tail than A4, indicating that standardization does not fully harmonize higher-order distributional properties. Figure 7D summarizes these raw differences, highlighting a large shift for p-tau217 and a smaller but consistent shift for Aβ42/40.

These findings demonstrate that plasma biomarker distributions differ systematically across cohorts in ways that are not fully corrected by standard preprocessing. Such distributional mismatches are consistent with dataset shift and provide a mechanistic explanation for the calibration errors observed in Figure 6. Because machine learning models learn decision boundaries based on cohort-specific feature distributions, shifts in mean, variance, and shape can distort how those boundaries apply to new data. This effect is particularly relevant for p-tau217, which was the dominant predictive feature across models. Together, these results indicate that assay-related and cohort-specific biomarker differences are a key driver of miscalibration and of reduced NPV under cross-cohort deployment^17, 31, 45, 47, 48, 60^.

### Decision Curve Analysis: Net Clinical Benefit Under Cross-Cohort Deployment

Clinical utility was further evaluated using decision curve analysis (Figure 8), which compares net benefit across a range of threshold probabilities for within-cohort and cross-cohort model deployment^17, 43, 48^. Within-cohort models demonstrated consistently positive net benefit across clinically relevant thresholds in both ADNI and A4. In ADNI, internally validated models maintained higher net benefit than both “treat all” and “treat none” strategies over a broad threshold range, indicating meaningful clinical utility for identifying amyloid-positive individuals. Similarly, within-cohort A4 models showed sustained net benefit, although the magnitude was lower, consistent with the narrower disease spectrum and lower signal variability in the preclinical cohort.

In contrast, cross-cohort deployment resulted in a systematic reduction in net benefit. When ADNI-trained models were applied to A4, net benefit was consistently lower than the corresponding within-cohort model across nearly all threshold probabilities, with the largest reduction observed at intermediate thresholds. At higher thresholds, net benefit approached that of the “treat none” strategy, indicating diminished clinical usefulness. A similar pattern was observed for A4-trained models applied to ADNI, although attenuation was less pronounced in this direction. Cross-cohort models remained above the “treat none” line but showed consistently reduced net benefit relative to within-cohort performance, indicating only partial preservation of clinical utility.

These findings indicate that even when discrimination remains relatively stable under cross-cohort transfer, clinical decision performance, as quantified by net benefit, can decline substantially. The reduction in net benefit was consistent with the observed declines in NPV and calibration, reinforcing the conclusion that deployment-related shifts in probability estimates have direct consequences for clinical utility. Decision-analytic measures of clinical utility were therefore more sensitive to dataset shift than traditional discrimination metrics.

### Prevalence-Adjusted NPV Simulation

To isolate the contribution of disease prevalence to NPV degradation, Supplementary Figure S5 modeled NPV as a function of prevalence using sensitivity and specificity estimated at Youden-optimal thresholds^17, 42, 48^. NPV declined nonlinearly as prevalence increased. Cross-cohort curves were uniformly lower than within-cohort curves because of modest sensitivity attenuation under transfer. At observed ADNI and A4 prevalence levels, simulated NPV closely matched empirical results.

These findings show that both prevalence differences and calibration shifts contribute to reduced rule-out performance. Even small reductions in sensitivity can translate into clinically meaningful decreases in NPV when disease prevalence is high.

### Summary of Key Findings

Collectively, these results demonstrate that plasma biomarker–based models achieve strong within-cohort discrimination and robust centiloid prediction. Cross-cohort deployment produces only modest AUC reduction but greater attenuation in regression performance. Clinically actionable metrics, particularly NPV, are substantially more sensitive to dataset transfer than discrimination. Calibration shifts and biomarker distributional differences provide mechanistic explanations for performance degradation, and prevalence amplifies modest sensitivity changes into clinically meaningful reductions in rule-out performance. These findings underscore the importance of cross-cohort validation, explicit calibration assessment, and assay-consistent biomarker generation before real-world deployment of blood-based screening tools.

## Discussion

In this study, we evaluated the performance and portability of plasma biomarker-based machine learning (ML) models for amyloid PET prediction within and across two complementary cohorts, ADNI and A4. The results support five interrelated conclusions.

First, plasma biomarker-based ML models demonstrate strong within-cohort performance for both binary amyloid classification and continuous centiloid prediction. Discrimination approached or exceeded AUC values of 0.90 in ADNI and 0.87 in A4, consistent with prior reports of plasma p-tau217-based prediction^20, 21, 26, 52^. Model structure analyses further demonstrated that biological signal hierarchy is stable across cohorts and algorithms, with p-tau217 consistently emerging as the dominant predictor, followed by Aβ42/40 and APOE ε4 count. These findings reinforce the robustness of core amyloid-related plasma biomarkers and support their biological validity.

Second, cross-cohort deployment results in reproducible but modest attenuation in discrimination (absolute AUC reduction of ∼4-7%) and greater attenuation in continuous centiloid prediction. Importantly, feature ranking and directionality remain stable under transfer, indicating that cross-cohort degradation is not driven by reversal of biological associations. These findings are consistent with broader observations that predictive performance can degrade under dataset shift even when underlying signal remains valid^45, 47, 61^.

Third, and most clinically consequential, cross-cohort deployment substantially degrades clinically actionable metrics. Negative predictive value (NPV) declined by approximately 20 percentage points when ADNI-trained models were applied to the preclinical A4 population. This reduction is substantially larger than the observed AUC decline and underscores that discrimination alone does not capture deployment risk. Given current appropriate-use recommendations emphasizing high NPV for screening applications^17, 48^, this degradation has direct implications for clinical implementation.

Fourth, calibration analyses reveal systematic probability misestimation under cross-cohort transfer. Even when AUC remains acceptable, predicted probabilities deviate from observed event rates, directly affecting NPV. Threshold-dependent analyses demonstrate that this instability affects the entire operating characteristic surface rather than a single decision point, and decision curve analysis confirms reduced net clinical benefit under deployment. These findings are consistent with established literature demonstrating that models optimized for discrimination may exhibit unstable probability estimates under distributional shift^45, 47^.

Fifth, biomarker distributional differences and disease prevalence provide mechanistic explanations for these observations. Plasma p-tau217 and Aβ42/40 distributions differ between ADNI and A4 despite within-cohort standardization. Empirical head-to-head comparisons of plasma p-tau217 assays demonstrate that platform and calibration differences can produce measurable shifts in biomarker distributions^31, 48^, consistent with earlier evidence of assay variability in Alzheimer’s disease biomarker research^32^. Such distributional mismatches likely drive calibration shifts under cross-cohort transfer. In addition, prevalence-adjusted analyses demonstrate mathematically that even modest sensitivity differences, when combined with prevalence variation, can produce large NPV changes. Thus, NPV degradation under deployment is both structurally predictable and empirically observed.

### From Signal Strength to Deployment Fragility

The stability of feature contributions across cohorts indicates that the underlying biological signal is preserved. SHAP analyses (Supplementary figure 3) demonstrate monotonic and coherent predictor effects, and feature importance rankings confirm consistent predictor hierarchy across algorithms. These findings suggest that the models capture genuine pathophysiological relationships rather than cohort-specific artifacts.

However, biological robustness does not guarantee deployment robustness. Cross-cohort attenuation in AUC, although modest, becomes clinically amplified when translated into predictive values. This divergence between discrimination and predictive value highlights a key translational insight: models may maintain ranking performance while losing decision reliability. This phenomenon is well described in the broader ML literature, where dataset shift can degrade probability calibration even when discrimination remains stable^47^.

### Calibration as the Critical Translational Axis

Our results indicate that calibration, not discrimination, is the principal axis of translational fragility. Cross-cohort models exhibited systematic miscalibration, shifting predicted probabilities relative to observed risk. Because NPV depends directly on predicted probabilities and prevalence, even small calibration deviations propagate into substantial changes in rule-out performance.

This finding aligns with prior evidence that modern ML models frequently exhibit miscalibration under domain shift^45^, and that standard recalibration approaches such as Platt scaling or isotonic regression may only partially mitigate these effects^47^. In our analyses, simple intercept recalibration improved performance modestly but did not eliminate direction-dependent degradation, suggesting that deeper distributional differences contribute to instability.

### Distributional Shift and Assay Heterogeneity

Distributional analyses reveal that plasma biomarkers, particularly p-tau217, exhibit cohort-level differences in mean, variance, and distributional shape. Although z-scoring aligns cohort means, higher-order differences persist, altering the effective operating region of learned decision boundaries.

These findings support the hypothesis that assay heterogeneity, including platform differences and calibration strategies, is a primary contributor to cross-cohort instability. Prior studies have documented inter-platform variability in AD biomarker measurement^32, 34, 62, 63^, and dataset shift theory predicts that such differences can degrade predictive performance under transfer^47^. The preservation of predictor directionality across cohorts suggests that biological relationships remain intact, whereas instability arises from measurement context rather than biological reversal.

### Prevalence Amplifies Deployment Effects

The prevalence-adjusted analyses provide a formal explanation for observed degradation. Because NPV is inversely related to disease prevalence and directly related to sensitivity, modest reductions in sensitivity under cross-cohort transfer are amplified in moderate-to-high prevalence populations. The close agreement between simulated and empirical NPV values supports this interpretation.

These findings emphasize a key translational principle: predictive value is not an intrinsic property of a model but a function of sensitivity, specificity, and population prevalence. Consequently, deployment into populations with different base rates can materially alter rule-out performance even when discrimination remains similar.

### Implications for Longitudinal Biomarker Modeling

Recent plasma p-tau217 “clock” models have demonstrated promise for staging disease progression within harmonized cohorts^28^. Our findings suggest that longitudinal models may face similar portability challenges if assay consistency and calibration stability are not maintained across deployment contexts. Thus, cross-cohort validation and harmonized measurement frameworks are likely prerequisites not only for screening classifiers but also for reliable implementation of longitudinal staging models.

### Clinical and Translational Implications

Taken together, our results demonstrate that plasma biomarker-based ML models are biologically robust yet deployment-sensitive. Clinically actionable metrics such as NPV are substantially more vulnerable to dataset shift than AUC. For screening applications in asymptomatic populations, high NPV and stable calibration are essential. Accordingly, cross-cohort validation should be considered a prerequisite for deployment, as external validation is a critical but often underperformed step in clinical prediction modeling^64, 65^. In addition, calibration assessment must accompany discrimination reporting, as both are required to evaluate clinical usefulness of prediction models^43, 66^. Finally, assay-consistent biomarker generation may represent the most effective translational lever, as variability in data generation and model validation remains a key barrier to safe clinical implementation of AI-based prediction tools^67^.

## Conclusions

Plasma biomarker–based ML models demonstrate strong within-cohort performance but exhibit reproducible, direction-dependent degradation when deployed across cohorts. While discrimination declines modestly, clinically actionable rule-out performance can decline substantially due to calibration shifts, distributional mismatch, and prevalence sensitivity. These findings underscore the importance of deployment-aware validation, calibration monitoring, and assay harmonization in translating plasma biomarkers into scalable clinical screening tools.

## Author Contributions Statement

Apurva Korni (ML data and figure generation and analyses, review of the manuscript), Ebrahim Zandi (Design and Conceptualization; Investigation; Supervision; Original ML model generation and Formal analysis; Writing-original and final draft ; Writing-Review & Editing; Resources; Visualization; Project Administration)

## ACKNOWLEDMENTS

Data used in the preparation of this article were obtained from the Alzheimer’s Disease Neuroimaging Initiative (ADNI) database (adni.loni.usc.edu). ADNI was launched in 2003 as a public-private partnership led by Principal Investigator Michael W. Weiner, MD. ADNI is funded by the National Institute on Aging, the National Institute of Biomedical Imaging and Bioengineering, and through contributions from industry partners. The grantee organization is the Northern California Institute for Research and Education, and the study is coordinated by the Alzheimer’s Therapeutic Research Institute at the University of Southern California. ADNI data are disseminated by the Laboratory for Neuro Imaging at the University of Southern California.

Data were also obtained from the Anti-Amyloid Treatment in Asymptomatic Alzheimer’s Disease (A4) Study. The A4 Study is funded by the National Institute on Aging, Eli Lilly and Company, the Alzheimer’s Association, and additional philanthropic contributors, and is led by investigators at Brigham and Women’s Hospital, Harvard Medical School, and the Alzheimer’s Therapeutic Research Institute at the University of Southern California.

## Data Availability Statement

The data supporting the findings of this study are available on request from the corresponding author.

## Abbreviation Full term

AD: Alzheimer’s disease
ADNI: Alzheimer’s Disease Neuroimaging Initiative
APOE ε4: Apolipoprotein E ε4 allele
A4: Anti-Amyloid Treatment in Asymptomatic Alzheimer’s Disease
Aβ42/40: Plasma amyloid-beta 42/40 ratio
AUC: Area under the receiver operating characteristic curve
BBMs: Blood-based biomarkers
CSF: Cerebrospinal fluid
GFAP: Glial fibrillary acidic protein
LightGBM: Light Gradient Boosting Machine
ML: Machine learning
MCI: Mild cognitive impairment
NfL: Neurofilament light chain
NPV: Negative predictive value
p-tau217: Plasma phosphorylated tau 217
PET: Positron emission tomography
PPV: Positive predictive value
PBBMs: Plasma-based biomarkers
R^2^: Coefficient of determination
RMSE: Root mean square error
ROC: Receiver operating characteristic
SHAP: SHapley Additive exPlanations
SVM: Support vector machine
XGBoost: Extreme Gradient Boosting

---

**Supplementary Table S1.**
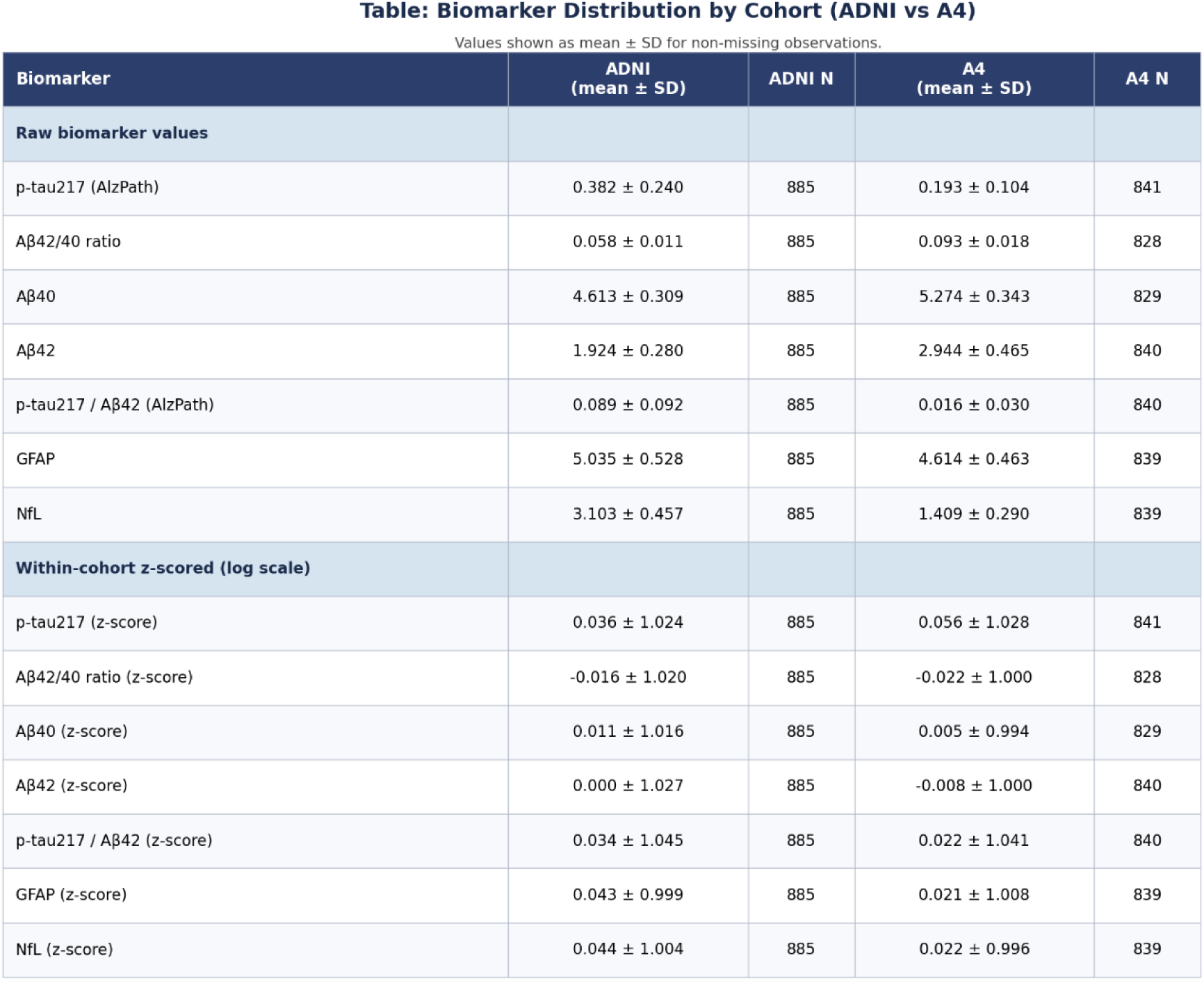
Means and variances for primary biomarkers.

| Biomarker | ADNI<br>(mean ± SD) | ADNI N | A4<br>(mean ± SD) | A4 N |
| --- | --- | --- | --- | --- |
| **Raw biomarker values** |  |  |  |  |
| p-tau217 (AlzPath) | 0.382 ± 0.240 | 885 | 0.193 ± 0.104 | 841 |
| Aβ42/40 ratio | 0.058 ± 0.011 | 885 | 0.093 ± 0.018 | 828 |
| Aβ40 | 4.613 ± 0.309 | 885 | 5.274 ± 0.343 | 829 |
| Aβ42 | 1.924 ± 0.280 | 885 | 2.944 ± 0.465 | 840 |
| p-tau217 / Aβ42 (AlzPath) | 0.089 ± 0.092 | 885 | 0.016 ± 0.030 | 840 |
| GFAP | 5.035 ± 0.528 | 885 | 4.614 ± 0.463 | 839 |
| NfL | 3.103 ± 0.457 | 885 | 1.409 ± 0.290 | 839 |
| **Within-cohort z-scored (log scale)** |  |  |  |  |
| p-tau217 (z-score) | 0.036 ± 1.024 | 885 | 0.056 ± 1.028 | 841 |
| Aβ42/40 ratio (z-score) | -0.016 ± 1.020 | 885 | -0.022 ± 1.000 | 828 |
| Aβ40 (z-score) | 0.011 ± 1.016 | 885 | 0.005 ± 0.994 | 829 |
| Aβ42 (z-score) | 0.000 ± 1.027 | 885 | -0.008 ± 1.000 | 840 |
| p-tau217 / Aβ42 (z-score) | 0.034 ± 1.045 | 885 | 0.022 ± 1.041 | 840 |
| GFAP (z-score) | 0.043 ± 0.999 | 885 | 0.021 ± 1.008 | 839 |
| NfL (z-score) | 0.044 ± 1.004 | 885 | 0.022 ± 0.996 | 839 |

**Supplementary Figure S1.**
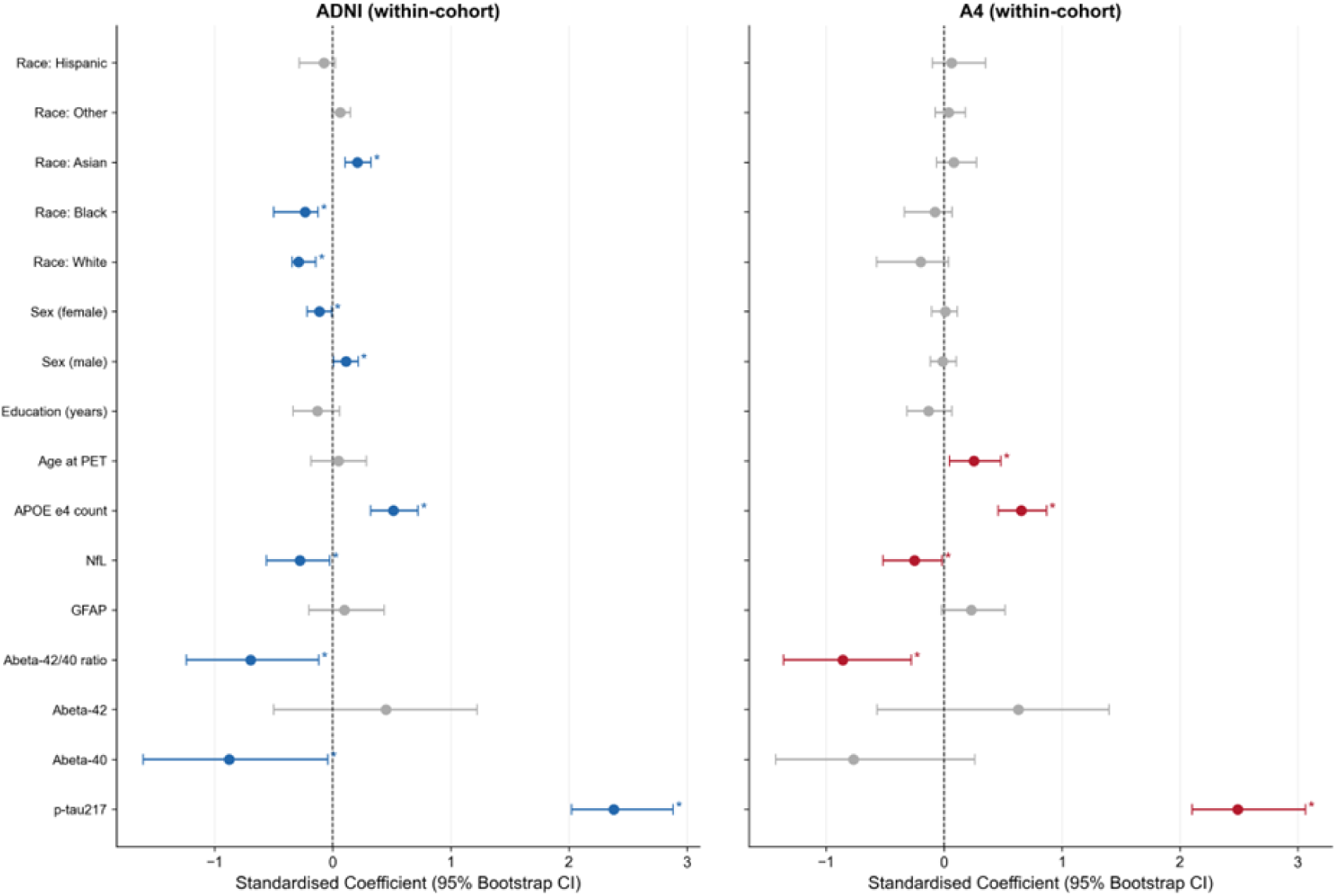
Standardized logistic regression coefficients for within-cohort amyloid PET classification models. Forest plots display standardized regression coefficients (with 95% bootstrap confidence intervals) from logistic regression models predicting amyloid PET positivity within ADNI (left panel) and A4 (right panel). All predictors were z-scored within cohort prior to model fitting. Points represent standardized log-odds coefficients; horizontal lines indicate 95% bootstrap confidence intervals. The vertical dashed line indicates a coefficient of zero. Asterisks (*) denote coefficients for which the 95% confidence interval does not cross zero. Predictors include demographic variables (age, sex, education, race), genetic risk (APOE ε4 allele count), and plasma biomarkers (p-tau217, Aβ40, Aβ42, Aβ42/40 ratio, GFAP, NfL). Separate within-cohort models were estimated for ADNI and A4. Standardized coefficients represent:

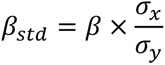 Because predictors were z-scored, coefficient magnitude directly reflects relative log-odds contribution per standard deviation change in predictor. Bootstrap confidence intervals were used rather than asymptotic Wald intervals to provide more robust uncertainty estimates.

**Supplementary Figure S2.**
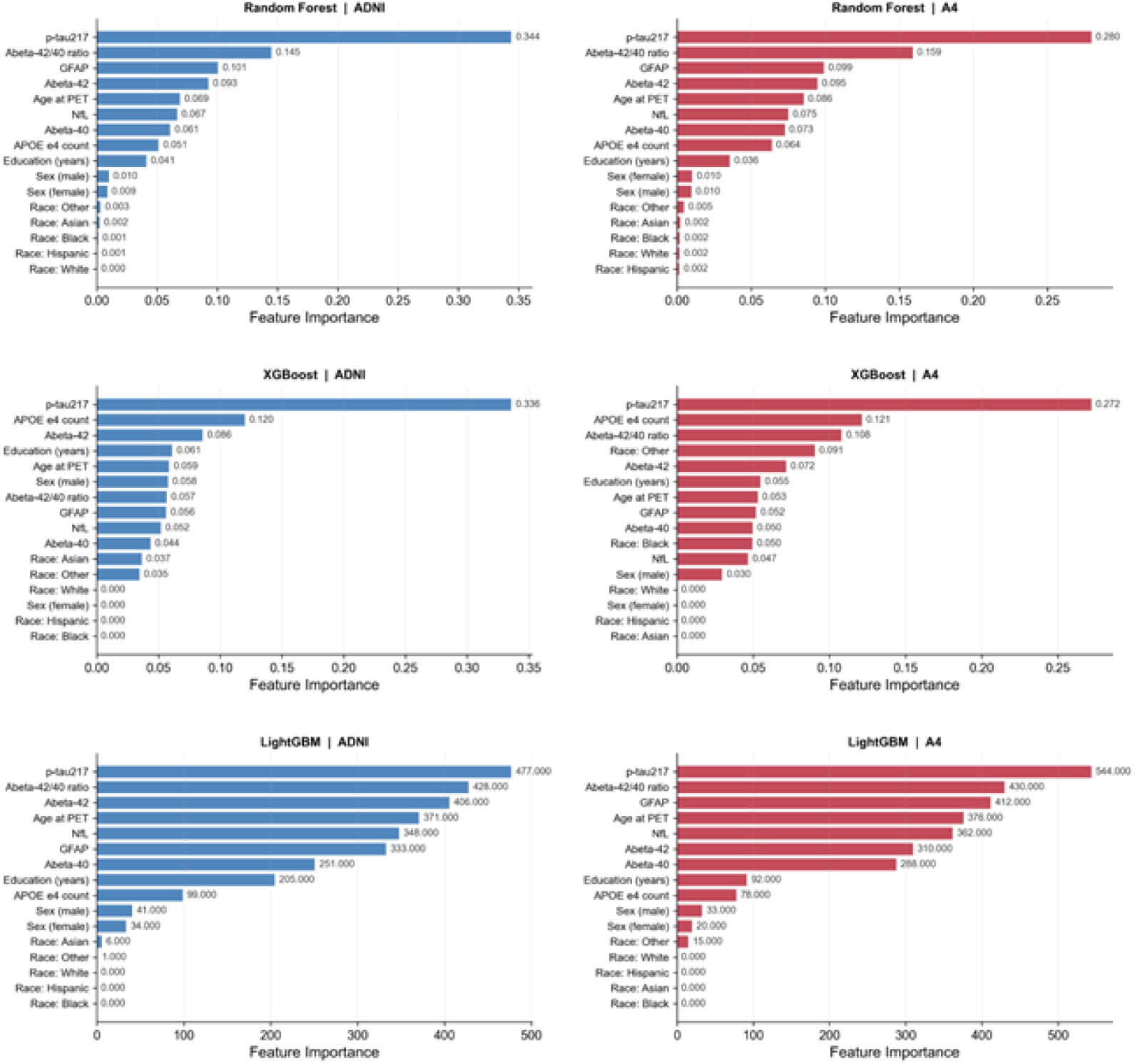
Feature importance for amyloid PET classification across machine learning algorithms and cohorts. Bar plots display feature importance rankings for amyloid PET classification models trained within ADNI (left panels, blue) and A4 (right panels, red). Feature importance is shown for three algorithms: Random Forest (top row), XGBoost (middle row), and LightGBM (bottom row). For tree-based models (Random Forest and XGBoost), feature importance values represent normalized mean decrease in impurity. For LightGBM, importance values reflect total gain contribution across trees. Predictors include plasma biomarkers (p-tau217, Aβ42, Aβ40, Aβ42/40 ratio, GFAP, NfL), demographic variables (age at PET, sex, education, race), and genetic risk (APOE ε4 allele count). Bars are ordered by decreasing importance within each model. Tree-based feature importance measures: Random Forest: Mean decrease in impurity (Gini importance), XGBoost/LightGBM: Gain contribution across boosting iterations Because importance is algorithm-specific, absolute values are not directly comparable across models; ranking consistency is the key interpretive metric.

**Supplementary Figure S3.**
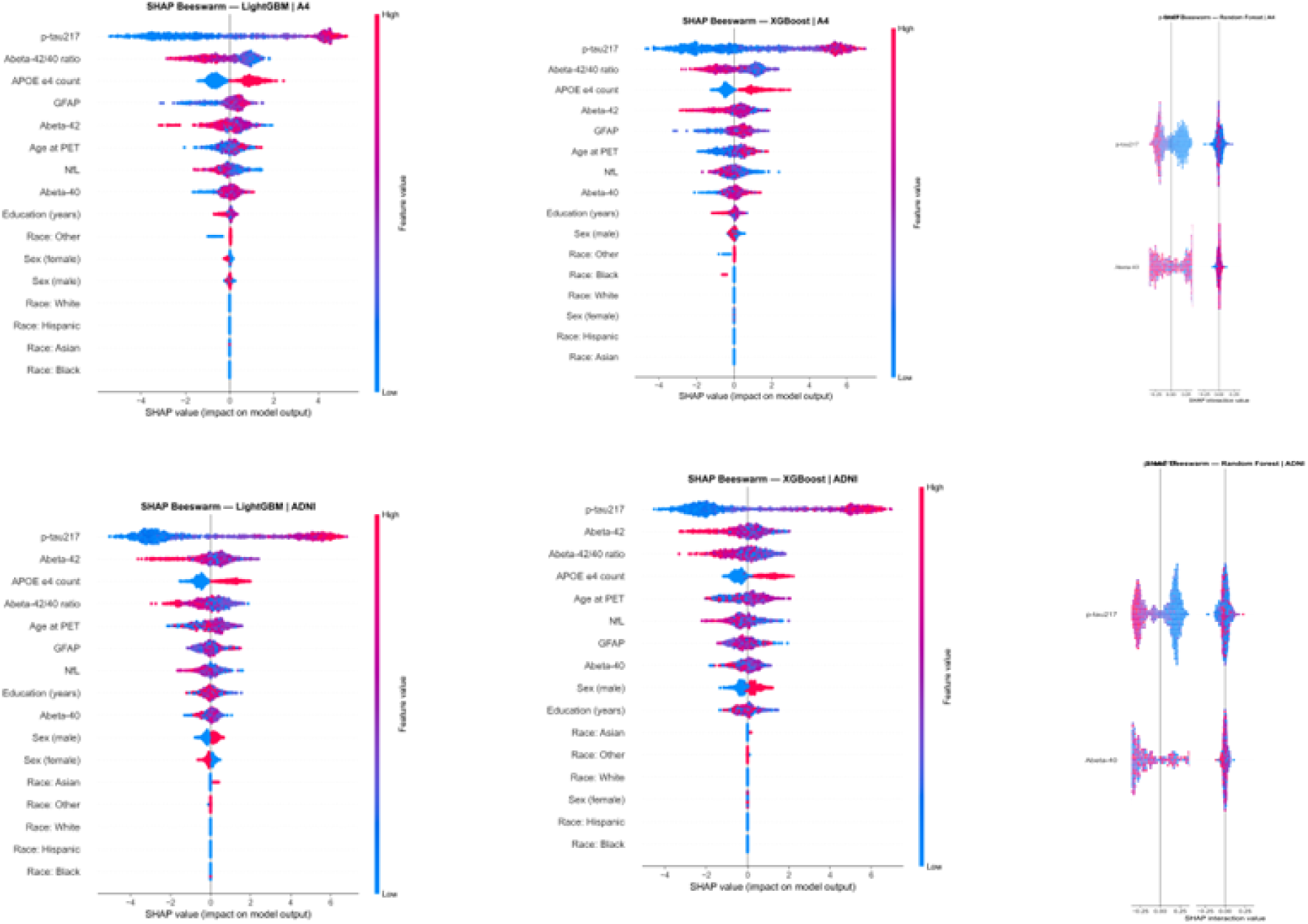
SHAP beeswarm plots illustrating feature contributions to amyloid PET classification across cohorts and algorithms. SHAP (SHapley Additive exPlanations) beeswarm plots display feature-level contributions to model predictions for amyloid PET status in A4 (top panels) and ADNI (bottom panels). Models include LightGBM, Random Forest, and XGBoost. Each point represents one participant. The x-axis indicates the SHAP value (impact on model output), with positive values increasing predicted probability of amyloid positivity and negative values decreasing it. Features are ordered by mean absolute SHAP value. Point color reflects feature value (blue = low, red = high). Vertical gray lines denote zero contribution. Wider horizontal dispersion reflects greater impact on model prediction. SHAP values are based on Shapley values from cooperative game theory and provide:Local explanation (per individual prediction), additive feature attribution, model-agnostic interpretation For tree-based models, TreeSHAP provides exact SHAP values efficiently. SHAP magnitude reflects contribution to log-odds prediction.

**Supplementary Figure S4.**
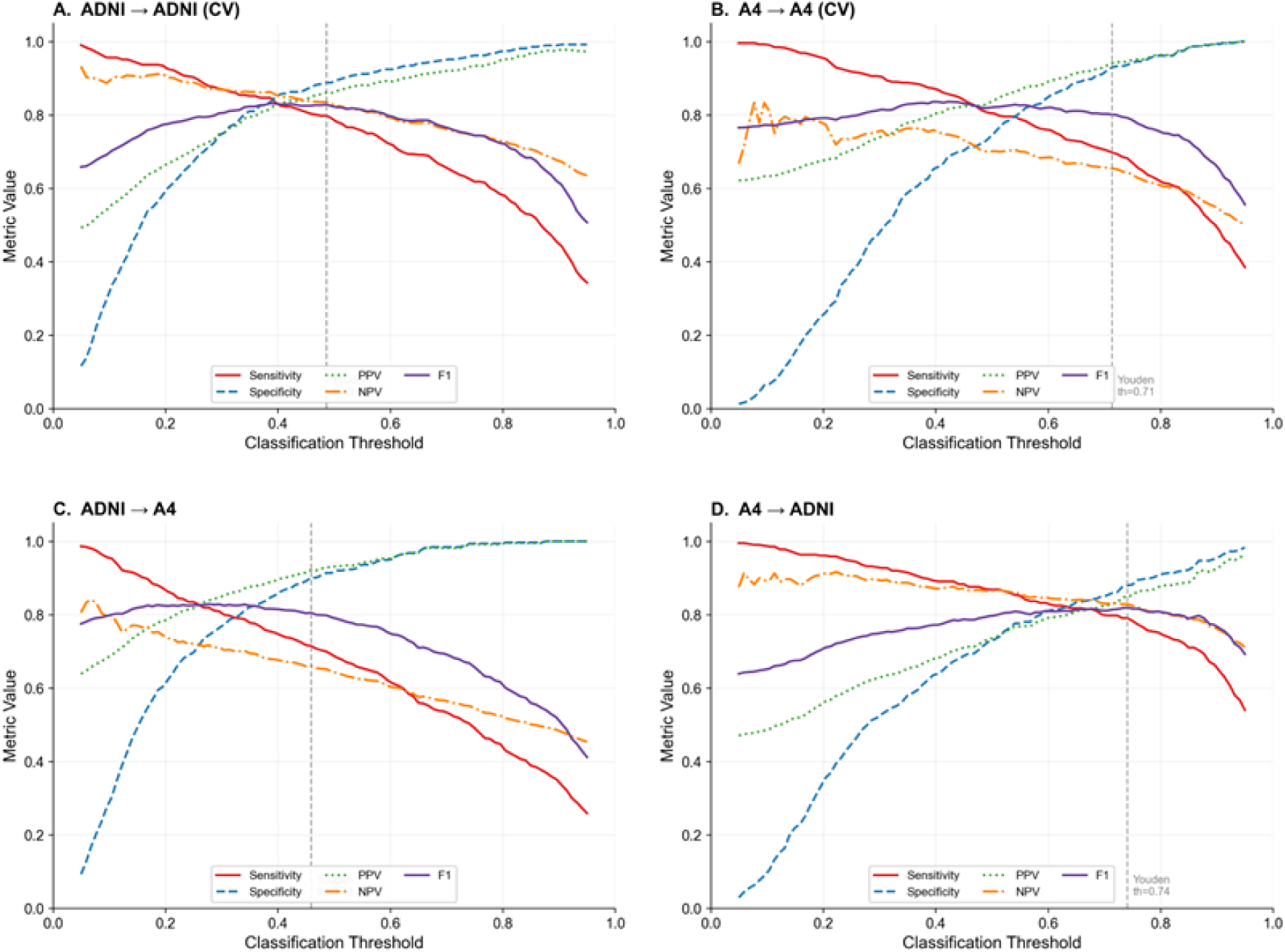
Sensitivity, specificity, positive predictive value (PPV), negative predictive value (NPV), and F1 score across classification thresholds for logistic regression models. Performance metrics are shown as a function of classification probability threshold for logistic regression models predicting amyloid PET status. Panels represent: **A. ADNI → ADNI (internal cross-validation)**, **B. A4 → A4 (internal cross-validation)**, **C. ADNI → A4 (cross-cohort transfer)**, **D. A4 → ADNI (cross-cohort transfer)** Curves depict sensitivity (solid red), specificity (blue dashed), PPV (green dotted), NPV (orange dash-dot), and F1 score (purple). Vertical dashed lines indicate the Youden-optimal threshold within each training cohort. Metric values were calculated across the full range of probability thresholds (0–1). Confidence intervals were estimated using bootstrap resampling.

**Supplementary Figure S5.**
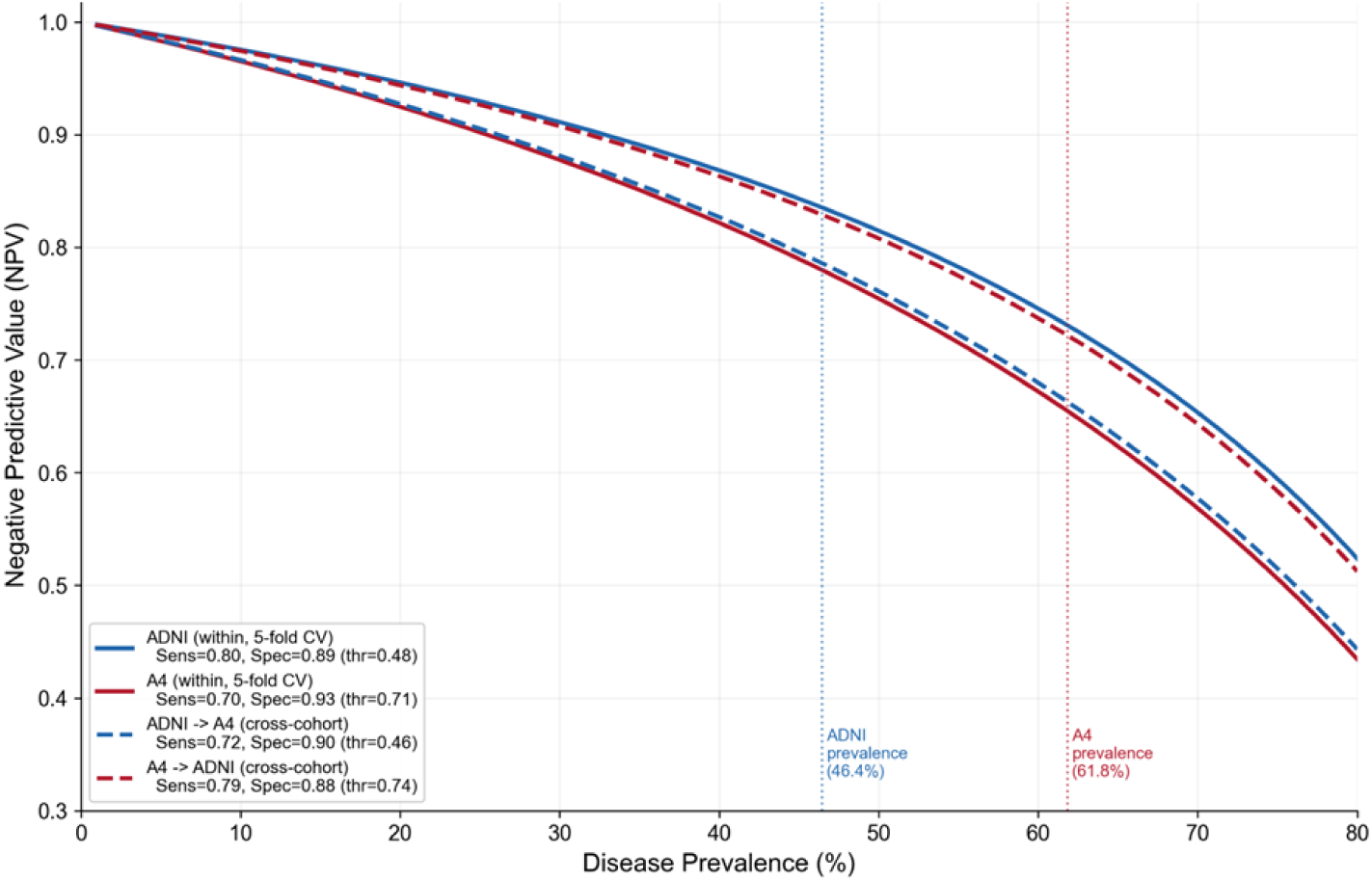
Prevalence-adjusted negative predictive value (NPV) simulation for logistic regression models. NPV is plotted as a function of disease prevalence using sensitivity and specificity estimates derived from logistic regression models at Youden-optimal thresholds. Solid lines represent within-cohort cross-validated performance (ADNI in blue, A4 in red). Dashed lines represent cross-cohort deployment (ADNI→A4 in blue dashed; A4→ADNI in red dashed). Sensitivity and specificity values used for each curve are shown in the legend. Vertical dotted lines indicate observed amyloid prevalence in ADNI (blue; 46.4%) and A4 (red; 61.8%). NPV was calculated using the standard prevalence-adjusted formula:

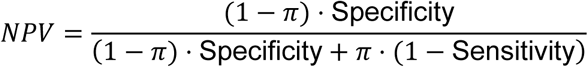 where *π* represents disease prevalence.

